# Multiplexed visual test with readout synchronized to each respective clinical threshold for rogue anti-cytokine autoantibodies

**DOI:** 10.64898/2026.08.02.26359144

**Authors:** Houda Shafique, Stéphane Bernier, Andy Ng, Lucie Roussel, Donald C. Vinh, David Juncker

## Abstract

Rapid tests with visual readout can quickly help identify and triage at-risk patients. However, multiplexed visual tests (MVTs) for targets with clinical thresholds above the limit of detection — e.g., rogue autoantibodies (raAbs) — are lacking because test interdependency and cross-reactivity make optimization intractable. Here, we introduce a conceptual and experimental framework to synchronize visual readout and clinical thresholds for multiple, cross-reacting targets simultaneously, and illustrate it with a 3D-printed, structurally-preprogrammed capillaric MVT for anti-interferon (IFN)-α and -ω raAbs and anti-SARS-CoV-2 spike protein (anti-SCoV2) antibodies. Using design of experiments, we sought and identified parameters that collectively govern the background of all tests (buffer composition, ionic strength), and ones that individually govern assay signal and sensitivity (capture probe density, sample volume), thus enabling both collective background reduction and independent tuning of test line visual threshold. The instrument-free MVT is highly sensitive (pg-ng mL^−1^), reproducible (CV<10% in plasma), and completed in <1 h. We benchmarked the threshold-calibrated MVTs to microplate ELISA with 41 COVID-19 patient plasma samples yielding ROC-AUCs of 0.97, 1.00, and 0.98 for anti-IFN-α, -IFN-ω, and -SCoV2 tests, respectively. The proposed framework for synchronizing multiple visual readouts with respective clinical thresholds, combined with capillarics, opens the door to instrumentation-free MVTs for point-of-care use.

## Introduction

Rapid tests in the form of lateral flow assays (LFAs) are based on functionalized nitrocellulose strips, are simple to use, offer visually interpretable yes/no results from test and control lines observed by naked eye, and are ubiquitous for at-home infectious disease (e.g., COVID-19, Influenza A/B) and pregnancy testing. Their sensitivity constitutes a limiting factor, and LFAs operate at their maximal sensitivity, all while recognizing that they suffer from false negatives for samples with low target concentration. Their overall simplicity including immediate visual readout, high specificity (i.e., low false positives), and short time-to-result underpin their popularity. These qualities prompted the development of multiplexed visual tests (MVTs) for combination infectious disease testing (with again sensitivity-limited performance) with 3 and even 4 targets on the same strip (e.g., Abbott Panbio COVID-19/Flu A&B panel self-test).^1^ Optimizing and improving MVTs remains an area of active research.^2^ However, it is important to recognize that MVTs so far have been restricted to sensitivity-limited tests (and primarily for infectious diseases) which are but a subset of possible tests, as discussed in the following paragraphs. In these sensitivity-limited cases, multiplexing is facilitated because susceptibility to cross-reactivity is greatly reduced when most targets are either not present or below the limit of detection (LOD) for any given sample and patient.

In diagnostics in general, the sensitivity of a test must be tuned to match the clinical application and the concentration range of the target being measured, which in many cases is well above the LOD of the test and ideally within the linear range where the analytical sensitivity and precision are highest. The clinical threshold thus guides the test design, and requires either quantitative readout based on signal amplitude or colorimetric analysis, and interpretation against calibrated standards. Common examples of simple tests include urine dipsticks (e.g., Roche Chemstrip, Siemens Multistix) that use printed colorimetric reference charts to estimate analyte levels of leukocytes, glucose, bilirubin, among others to monitor kidney disease, urinary tract infections, or diabetes at the point-of-care.^3,4^ Line immunoassays (e.g., Mikrogen recomLine, recomBlot) are used to semi-quantitatively assess panels of antibodies and allergens measured with test lines on nitrocellulose strips that are scanned on instruments often housed in a central facility, and compared against digital reference standards.^5,6^ In all these formats, interpretation depends on comparing visual intensity to an external standard, which can be locally interpreted by a clinician or digitally scanned for controlled readout and referenced against calibrated algorithms for thresholds, making them less suitable for point-of-care deployment compared to conventional LFAs. Yet, in many of these cases, a simple yes/no test precisely tuned to the clinical threshold of a given target may be adequate to help triage patients for treatment or reflex testing.^7^

There are many unmet needs for multiplexed simple yes/no readout at the point-of-care based on the appearance of a line without need for instrumentation, and there are no MVTs for non-sensitivity-limited scenarios. Indeed, when target concentrations are high, analytical sensitivity must be reduced to match a clinical threshold while maintaining individual specificity, and multiplexing becomes quickly intractable because assay interdependencies are generally poorly understood. Thus, while optimizing the visual threshold at desired detection thresholds for a single LFA is possible,^8,9^ rationally designing threshold-calibrated MVTs that simultaneously account for assay crosstalk, reagent cross-reactivity, and multiple interdependent, sometimes opposing, constraints,^10^ remains an open question.

Rogue autoantibodies (raAbs) against type I interferons (IFNs) can serve a risk factors for poor disease outcome and are thus a clinically relevant example of this challenge because different anti-type 1 IFN raAbs exist, they are detectable in healthy individuals, and hence the tests must be thresholded, and should each be detected individually, yet they cross-react.^11^ Anti-type 1 IFN raAbs have important clinical implications: raAb prevalence increases with age, and elevated levels of raAbs neutralizing type I IFNs, primarily IFN-α and -ω, are detected in ∼10% of patients with severe COVID-19,^12–14^ affecting 15-20% of critical cases overall and >20% of those aged >70 years,^12,15^ with >90% occurring in senior male patients.^12^ Recombinant type I IFNs not targeted by these raAbs, such as IFN-β, may restore antiviral immunity, thus requiring distinction between different anti-IFN raAbs. Similar outcomes have been reported in other viral infections.^16–18^ Screening for anti-type I IFN raAbs, alongside anti-SARS-CoV-2 antibodies (commonly against the spike protein, [anti-SCoV2]),^11,19^ was recognized for its potential for risk stratification and targeted interventions such as plasmablast depletion, plasmapheresis, and early antiviral therapy (e.g., nirmatrelvir/ritonavir [Paxlovid]). Yet despite their clinical significance and the need for timely multiplexed raAb detection, there are no diagnostic tests suitable for point-of-care screening.

raAbs are currently detected using laboratory-based solid-phase assays that require peripheral readers and depend on technical expertise. They include the enzyme-linked immunosorbent assay (ELISA), which is run in well plates and thus primarily limited to batch testing for the simultaneous analysis of many samples, but only detecting single antibodies.^20,21^ Multiplex detection of raAbs for disease profiling is possible using bead-based immunofluorescence assays and protein microarrays (e.g., MILLIPLEX, BioPlex 2200, FIDIS Lumine× 100, AtheNA Multi-Lyte) with 15-128-plex detection of anti-cytokine, autoimmune, and cancer autoantibodies.^6^ Similarly, line immunoassays are commonly used but readout is by imaging with a peripheral analyzer and comparing line intensities to digital reference standards. These assays suffer from high background signals because their protein-adsorbing surfaces promote non-specific binding (NSB) of human IgG from patient plasma,^22,23^ and are susceptible to cross-reactivity in multiplex detection.^24^

We recently introduced 3D-printed microfluidic chain reaction^25^ that expand the repertoire of structurally-preprogrammed microfluidic capillaric circuits (CCs), and made the ELISA-chip,^26^ which is a self-contained CC that structurally encodes and automates the liquid handling algorithm of an ELISA using the capillary phenomena only. It thus circumvents the need for peripherals and external power. Capillarics and ELISA-chips can directly be manufactured by photopolymerization 3D printing using hydrophilic inks,^27^ including with very low-cost (US$150-600) liquid crystal display (LCD) 3D printers.^28^ ELISA-chips with nitrocellulose strip as assay substrate have been used for single-plex detection of SARS-CoV-2 antibodies and antigen, as well as for IFN-γ, and were optimized for high sensitivity with both visual readout and imaging for quantification.^25,26,28^ ELISA-chip tests take longer than LFAs, but they have better sensitivity, reduced backgrounds, and improved reproducibility, notably thanks to programmable multi-step assays with washing steps that help mitigate NSB.

Here we present a conceptual and experimental framework for designing MVTs with visual readout synchronized to respective clinical thresholds for multiple cross-reacting targets simultaneously, and demonstrate it with a 3D-printed capillaric serotest for raAbs against two type 1 IFNs and anti-SCoV2 antibodies in clinical plasma. Our approach relies on a conceptual framework that separates assay optimization into collective parameters that govern background suppression and individual parameters that independently tune the visual threshold of each antibody. We implement this strategy using statistical design of experiments (DoE)^29–31^ to systematically and quantitatively seek and identify both parameter types, and use them to first collectively reduce the background across the strip, then independently tune test line visual threshold. While DoE is commonly used for assay optimization,^32,33^ particularly to reduce assay sensitivity,^2^ we use it for key parameter identification to tune our assay performance and for assessing interdependencies to enable rational design of threshold-calibrated capillaric MVTs. To illustrate the clinical potential, we validated 3D-printed capillaric MVTs in a 41-patient cohort by benchmarking patient stratification of the visual readout against conventional microplate ELISA, observing excellent concordance.

## Results

The concept of designing capillaric MVTs, according to template of the ELISA-chip,^25,26,28^ with readout synchronized to each respective antibody clinical threshold is summarized in **Figure 1**. The conceptual and experimental framework separates assay optimization into tuning key individual assay controlling parameters and collective background reduction parameters, and using statistical DoE to seek, identify, and quantify both parameter types used for downstream optimizations, **Figure 1a**. This framework allows for tuning of each assay’s visual readout to clinically meaningful cutoffs such that the visual ON state appears only when the antibody concentration exceeds the clinical positivity threshold, **Figure 1b**. This enables independent tuning of each assay while maintaining a unified visual readout on a single test strip to achieve threshold-calibrated MVTs, **Figure 1c**. The clinical need to identify patients with anti-IFN-α and -ω raAbs in the context of COVID-19 infection highlights the utility of threshold-calibrated MVTs for risk stratification, **Figure 1d**, and the design of an associated capillaric MVT for triplex antibody detection in patient plasma is shown in **Figure 1e-f**. Finally, we validated visual readout for patient stratification and benchmarked performance against microplate ELISA showing potential for clinical use, **Figure 1g**.

**Figure 1.**
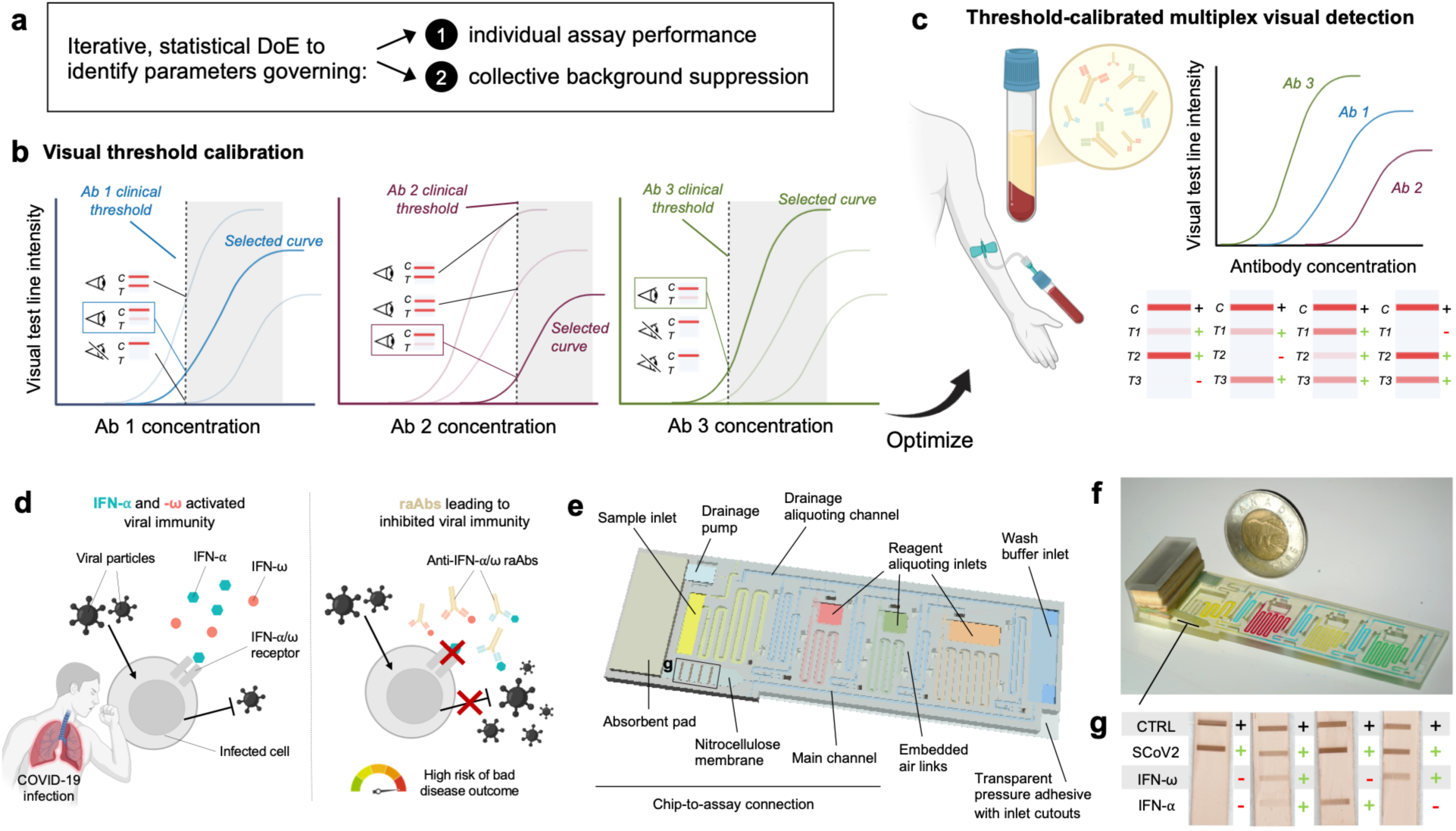
Overview of capillaric multiplexed visual tests (MVTs) with readout synchronized to clinical positivity thresholds for anti-type 1 IFN raAbs and anti-SCoV2 antibodies. (a) Conceptual and experimental framework for designing threshold-calibrated MVTs; using iterative, statistical DoE enables the identification and distinction of both (1) individual assay parameters governing signal strength and analytical sensitivity, and (2) collective parameters controlling background suppression. (b) Visual threshold calibration. Each antibody assay binding curve is tuned so the visual ON readout aligns with the respective clinical positivity threshold, and the curve with the visual response at the clinical threshold is selected. Parameters governing curve tunability (individually and collectively) need to be sought and identified to synchronize thresholds. “C” labels assay control lines, and “T” labels assay test lines. (c) Using DoE-guided key parameter identification to synchronize visual readout with clinical thresholds, and considering assay crosstalk, threshold-calibrated MVTs were designed onto single test strips based on clinically meaningful cutoffs. (d) Risk stratification. The immune response to COVID-19 infection is mediated by type I IFNs-α and -ω that promote antiviral activity, but that can be suppressed by high titers of pre-existing raAbs, increasing the risk of severe disease outcome; high raAb levels require therapeutic intervention, whereas basal levels remain below the clinical threshold. (e) Capillaric MVTs that structurally encode aliquoting and sequential delivery of assay reagents to a nitrocellulose membrane patterned with capture antigens of interest. (f) Image of 3D-printed capillaric MVTs with a Canadian $2 coin; microchannels are filled with food dyed water for visualization. (g) Visual readouts for anti-IFN-α and -ω raAbs and anti-SCoV2 test lines are synchronized with clinical thresholds for each target.

### Identification of key individual parameters governing assay LOD, linear range, and visual threshold, and assay interdependency

Experiments performed using statistical DoE follow the steps shown in **Figure 2a**. Briefly, for Taguchi orthogonal arrays, the example shown graphically varies 4 factors (A, B, C, and D) over 3 levels (1, 2, and 3) for a total of 9 experiments, i.e., Taguchi L_9_(3^4^), sampling an experimental space comprising of 3^4^ = 81 combinations; details about the breakdown, symmetric sampling, and nomenclature have been described elsewhere.^29–33^ Taguchi DoE provides quantitative estimation of the main effects and some two-way interactions while minimizing statistical confounding.^29,34,35^ To better parse the parameter space, the assay can be investigated at points where there are measurable changes to the signal in response to perturbation; this includes both near the LOD and in the linear range. In quantitative assays, focus is typically placed on variability near the detection limit; however, the inherently low signal in this regime can obscure the influence of certain experimental factors and may not generate enough non-zero data points. Performing dual measurements addresses this limitation and (1) distinguishes parameter effects at different points of the binding curve, and (2) determines whether the same parameters govern signal strength under low-signal, flat-curve conditions and high-signal, steep binding-curve conditions. Sampling two binding curve points therefore yields more informative and broadly translatable insights to guide subsequent optimization. Orthogonal array symmetry allows parameter effects and interactions to be isolated by averaging grouped runs and quantified using analysis of variance (ANOVA) to determine the effect size and relative contribution, i.e., “weight”, see Supplementary **Note S1** for the calculation details.

**Figure 2.**
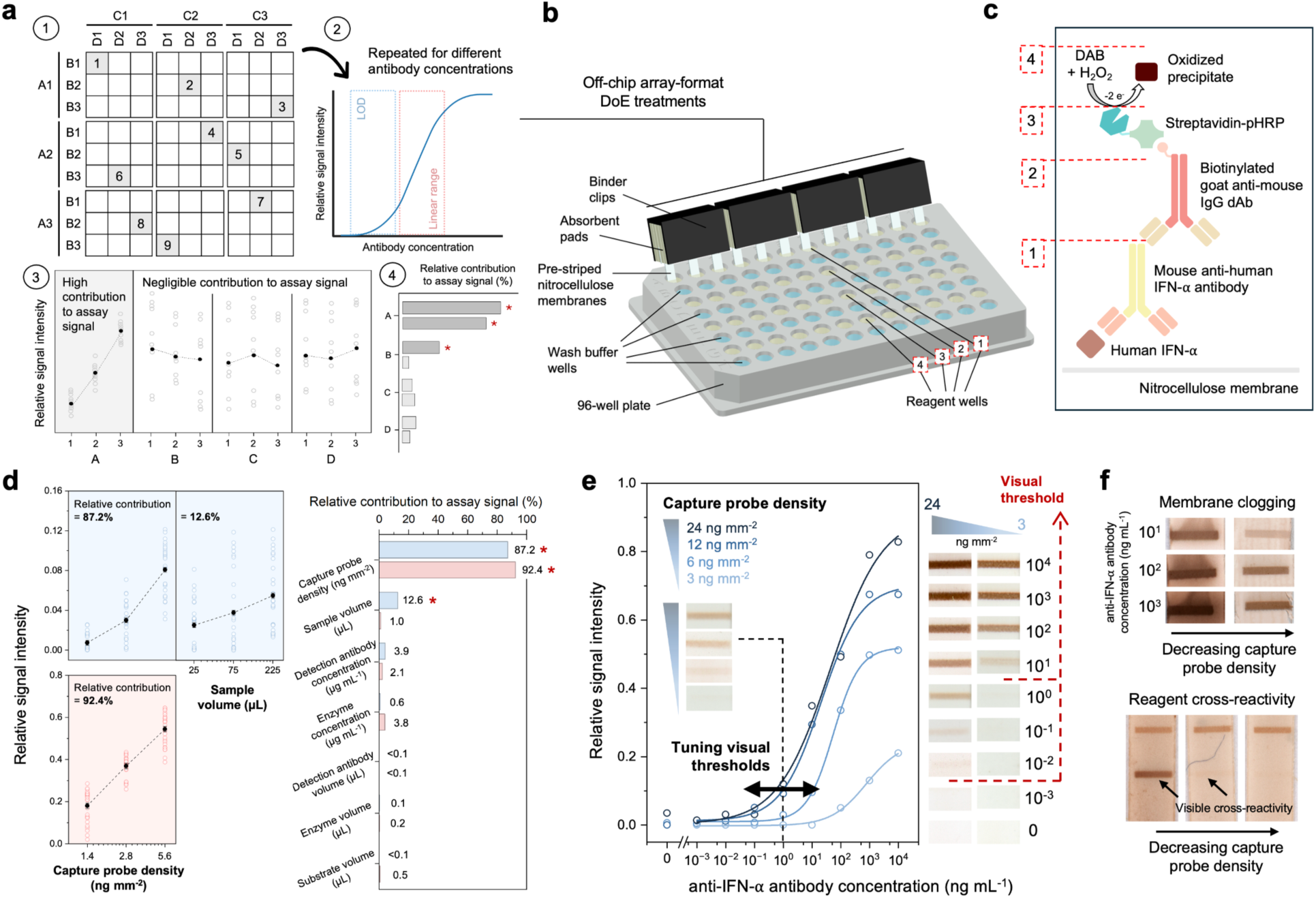
Statistical DoE to identify and quantify key individual parameters governing assay signal strength and analytical sensitivity. (a) Schematic of the DoE workflow, (1) testing 4 factors (A-D) at 3 levels (1-3) in 9 experiments, which (2) are replicated for analyte concentrations close to the LOD (blue outline) and in the linear range (red outline); (3) parameters, their interactions, and their relative contributions are individually quantified to assess impact on assay response, then (4) ranked to isolate key parameters driving performance. (b) Off-chip array-format experiments testing 12 DoE conditions in parallel with a well plate. (c) ELISA workflow for the detection of a mouse anti-human-IFN-α. (d) DoE response graphs close to the LOD (top) and in the linear range (bottom), and Pareto chart of ranked contribution to the assay signal uncover capture probe density as the dominating factor, while sample volume contributes only close to the LOD (identified by a red *); other factors are negligible. Solid data points show the mean ± standard error across grouped runs; hollow data points show the results across all experimental runs. (e) Tuning of visual readout by adjusting capture probe density. Inset shows signals for 1 ng mL^−1^ of mouse anti-human-IFN-α sample antibody, and right panel shows the visual threshold shifts ∼1000-fold with a difference in capture probe density from 24 to 3 ng mm^−2^. (f) Protein loading capacity, membrane clogging and reagent cross-reactivity govern the upper limit of visual detection.

Initial experiments were conducted in an off-chip array-format to increase throughput with 12 simultaneous experiments completed using hand-held, clamped nitrocellulose membranes in well plates, **Figure 2b**, see Materials and Methods for more details. The wells were loaded in the order of the assay reagents, including a biotinylated detection antibody, streptavidin-conjugated poly-horseradish peroxidase (pHRP) enzyme, and 3,3’-diaminobenzidine (DAB) as the enzyme substrate to generate a colorimetric readout after oxidation in the presence of hydrogen peroxide, producing a brown water-insoluble precipitate; each step was interspaced with a wash step, like a standard ELISA. Notably, in the absence of recombinant or isolated “*human* anti-human” IFN-α, a model immunoassay with *mouse* anti-human IFN-α was used, **Figure 2c**, as a surrogate target during key parameter identification.^36^

The assays were first established in buffer to identify key parameters governing performance, and colorimetric test line intensities following subtraction of adjacent up- and downstream background were measured for quantitative parameter identification.^26^ To start, reagent concentrations and volumes were varied. We examined seven factors across three different levels with a Taguchi L_27_(3^13^) orthogonal array, Supplementary **Table S1**, with ranges selected based on prior knowledge of the assay working ranges.^25,26^ Then, the table of conditions was run both near the LOD and in the linear range with sample antibody concentrations of 5 and 500 ng mL^−1^, respectively. The assay signals and response graphs for each parameter are given in Supplementary **Figure S1-2** and **Figure 2d**, respectively, and quantified effect size and relative contribution using ANOVA are given in Supplementary **Table S2**.

Near the LOD, the capture probe density and the sample volume had the highest relative contribution to the assay signal at 82.7% and 12.6%, respectively. In the linear range, the relative contribution shifted almost entirely to the capture probe density at 92.4% and variability attributed to changing the assay response became smaller for the sample volume at only 1.0%. These results indicate that capture probe density governs assay signal strength at both points, and the shift may suggest that the assay is in a capture probe-limiting regime, indicating that both the ON rate and maximal binding are reduced resulting in a lower signal. As probe availability increases, the ON rate increases, and higher signals can be obtained, and which near the LOD are increasing for larger sample volumes. Thus, thresholds for both capture probe density and sample volume need to be met to reach binding equilibrium, and strongly affect signal strength, while detection antibody, enzyme, and substrate concentrations and volumes, did not affect it in our context as they may already be provided in sufficient excess. DoE findings were validated as experiments showed that capture probe density could be used to modulate LOD and visual threshold, as well as shift and tune linear range according to the desired analytical sensitivity, **Figure 2e**. For example, an 8-fold difference in the capture probe density gave a ∼1000-fold difference in the visually detectable sample concentration.

The upper limit of capture probe density was set by either (1) streaking or clogging of the nitrocellulose membrane as a result of saturating the membrane protein binding capacity, or by (2) cross-reactivity between the antigen and other reagents, indicating that background signal from NSB could obscure weak signals or generate false positive test lines, **Figure 2f**. These results informed the choice of capture probe density, notably to mitigate potential cross-reactivity leading to assay interdependency, which we observed between IFN-ω test line and anti-IFN-α antibodies, Supplementary **Figure S4** (discussed later).

Next, we used DoE to evaluate whether washing steps impacted signal strength. We examined the four ELISA washing steps at two volumes, Supplementary **Table S3**. The nitrocellulose membrane backgrounds and response graphs for each wash step are given in Supplementary **Figure S3** and the relative contribution quantified in Supplementary **Table S4**. The wash step between the enzyme and substrate, was found to contribute highly to the assay response at 77.7%, which is in concordance with expectations and observations when the enzyme pre-mixes with the substrate, then begins to generate a colorimetric response before reaching the test line.

### Design of 3D-printed capillaric MVT

Capillaric MVTs were designed in accordance with DoE results and our previous work for CCs^25,26,37^ to encode the current assays. The nitrocellulose test strip was spotted with IFN-α, IFN-ω, SCoV2 test lines and a positive control line with biotinylated BSA (in this order in the flow direction). Based on the DoE experiments, we selected the sample volume to be 100 µL, the reagent volumes of the detection antibody and enzyme to be 35 µL (which was sufficient for reproducible results), the substrate to be 70 µL to provide sufficient reagent for up to four lines on the nitrocellulose membrane, and each of the four washing steps to be 25-35 µL of buffer. Based on our previous work, the volumetric precision of the 3D-printed chips was within 2-3%.^28^ The sample buffer was spiked with one at a time of each mouse anti-IFN-α, anti-IFN-ω, or anti-SCoV2 antibodies and the automated assay completed, **Figure 3a-b**, Supplementary **Video S1**.

**Figure 3.**
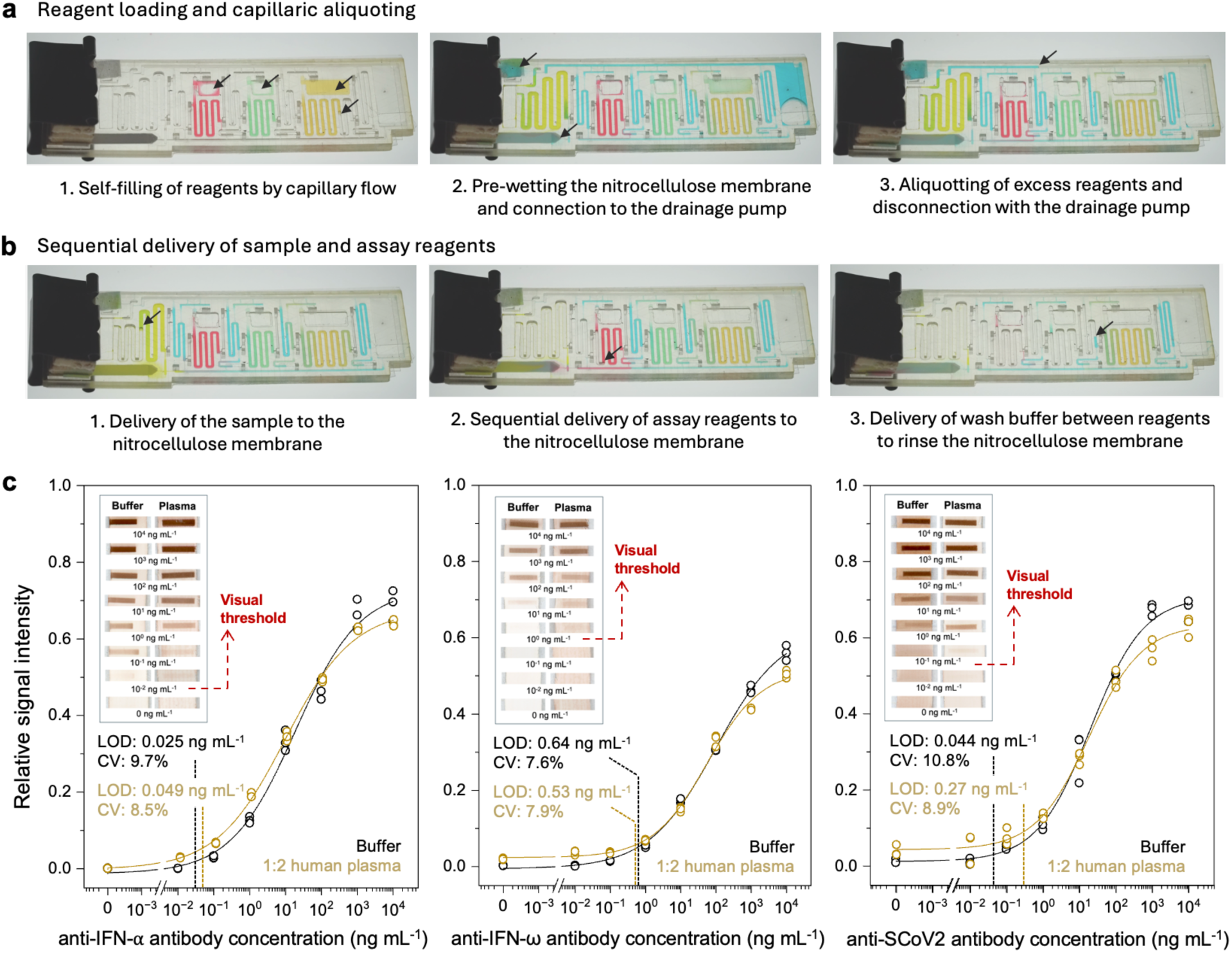
Capillaric MVT workflow and binding curves for the triplex visual detection of antibodies in buffer and plasma. (a) Capillaric MVTs with structurally encoded workflow of triplex visual detection on a 3D-printed preprogrammed CC shown with food dyed water running in the chip; reagent reservoirs are first metered to their correct volume and the excess is drained, followed by (b) sequential delivery of sample and assay reagents with wash steps in between to the pre-spotted nitrocellulose test strip. Black arrows are a guide to the eye. (c) Binding curves for the detection of mouse anti-IFN-α, anti-IFN-ω, and anti-SCoV2 antibodies in buffer and 1:2 human pooled plasma across triplicate chips. Inset shows the visual test line for each concentration point in buffer and plasma. Dashed red line shows the visual threshold, i.e., the lowest concentration where the test line becomes visibly apparent and detectable by naked eye. Solid lines show a 4-point logistic fit, and measured LODs with corresponding CVs are labelled with dotted lines for buffer and plasma.

We observed cross-reactivity of anti-IFN-α antibody with the IFN-ω test line, which needed to be considered when optimizing multiplexed assays, and was mitigated by reducing the IFN-ω antigen density from 56 to 14 ng mm^−2^ and resolved assay interdependency between these two targets, Supplementary **Figure S4**; no cross-reactivity was seen on the SCoV2 and IFN-α test lines, suggesting that they may be tuned independently. The capture probe density for the IFN-α and SCoV2 lines was just below the appearance of a faint line at zero sample concentration.^25^ Then LODs were measured quantitively using the test line colorimetric signal intensity, as described previously;^26^ details are given in Supplementary **Note S3**. In the triplex assay condition, spotting densities of 21, 14, and 11.2 ng mm^−2^ were empirically optimized for IFN-α, IFN-ω, and SCoV2, respectively, resulting in measured LODs of 0.025 (CV: 9.7%), 0.64 (CV: 7.6%), and 0.044 (CV: 10.8%) ng mL^−1^ in buffer, for anti-IFN-α, anti-IFN-ω, and anti-SCoV2 antibodies, respectively, **Figure 3c**.

To evaluate compatibility with human plasma, we used 1:2 plasma:buffer dilution and measured LODs of 0.049 (CV: 8.5%), 0.53 (CV: 7.9%), and 0.27 (CV: 8.9%) ng mL^−1^ for anti-IFN-α, anti-IFN-ω, and anti-SCoV2 antibodies, respectively, **Figure 3c**.

The visual threshold was defined as the lowest sample concentration that produced a test line observable by the naked eye. The threshold concentrations were 0.01, 1, and 0.1 ng mL^−1^ for anti-IFN-α, anti-IFN-ω, and anti-SCoV2 antibodies, respectively, **Figure 3c**. Altogether, the triplex capillaric MVT time-to-result was 55 min, featured pg-ng mL^−1^ LODs with CVs <10% in plasma for each antibody assay, and enabled visual, naked-eye readout in the same concentration ranges.

### Identification of key collective parameters for suppressing background in human plasma

We optimized capillaric MVT for human plasma with mouse antibodies, and next proceed to optimize with an anti-human IgG detection antibody. NSB to the immobilized antigens on the test line or the nitrocellulose membrane is a known issue,^38–41^ and was evaluated here using pre-pandemic citrated pooled human plasma negative for both raAbs and anti-SCoV2 antibodies. We initially observed high background on all three test lines. Three commonly known sources of NSB to either the antigen test lines or nitrocellulose membrane include NSB of (1) assay reagents like the detection antibody, enzyme-conjugate, or substrate, (2) sample matrix components, like plasma IgGs, or (3) recognition of blocking agents (e.g., anti-BSA antibodies). We already established that enzyme-conjugate and substrate do not contribute to NSB (**Figure 3c**), and thus evaluated NSB of the anti-human IgG detection antibody to the antigen and membrane, Supplementary **Figure S5**, and towards blocking agents, Supplementary **Figure S6**, but did not observe significant NSB. Consequently, we reasoned that the key contributors to NSB were likely hydrophobic or electrostatic binding of plasma components. We then optimized NSB by varying detergents and their concentrations (to suppress hydrophobic interactions), as well as salts and their concentrations (to supress electrostatic interactions) in the plasma diluent buffer using DoE to identify key collective parameters that drive background suppression on all three test lines, **Figure 4a-b**. We also observed that NSB was sensitive to plasma dilutions, Supplementary **Figure S7**, and not to capture probe density, so high density was retained to increase linear range and minimize false negatives (see **Figure 2e**). Here, the factors governing NSB and signal strength appeared to be independent, which was operationalized to minimize NSB without negatively impacting the tunability and visual thresholding of each target.

**Figure 4.**
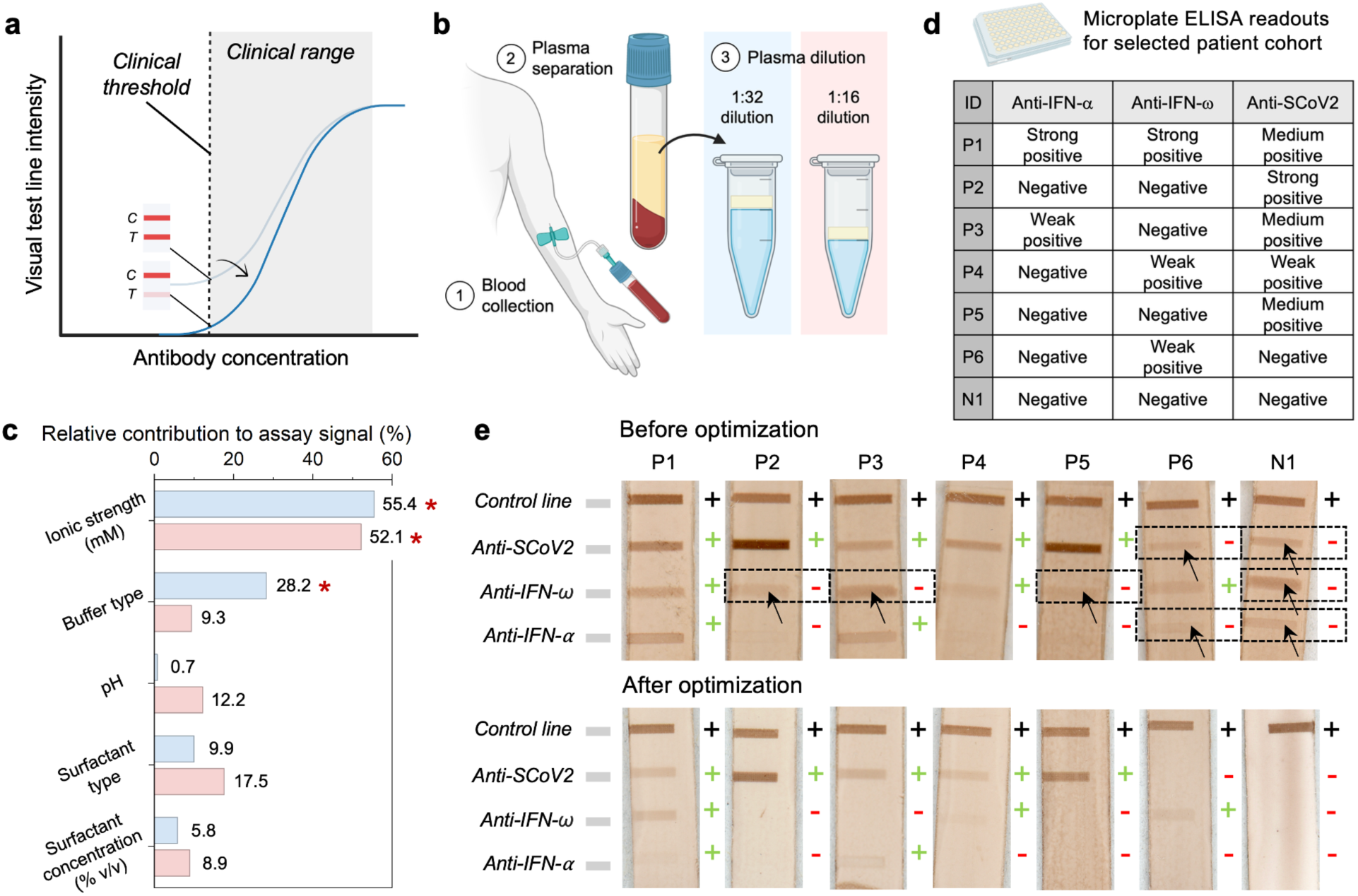
Identification of collective parameters controlling NSB and background suppression for visual readout in human plasma, and synchronization with clinical thresholds with patient plasma. (a) NSB signal lowers the visual threshold and must be considered when tuning visual readout to match the clinical positivity threshold. “C” labels assay control lines, and “T” labels assay test lines. (b) Two plasma dilutions were used to evaluate parameter effects on the NSB and background suppression. (c) DoE Pareto plots for relative contribution of buffer composition uncovering buffer ionic strength as major parameter affecting NSB, and buffer type (driven by Tris) only for 1:32 plasma dilution (identified by a red *); other factors were negligible. (d) Microplate ELISA binned readouts spanning different levels of positivity for different targets comprising six patient plasma positive controls (P1-6) and one pre-pandemic pooled plasma negative control (N1). (e) Capillaric MVT readouts for P1-6 and N1 before (top) and after (bottom) DoE-guided optimization to synchronize visual readout with each antibody’s clinical threshold. Black + symbols show the presence of a control line, and coloured +/− symbols show the expected result of the control plasma samples based on microplate ELISA as the reference method. Test lines in dashed black boxes show the presence of false positives with arrows identifying the visible test line.

Three sequential Taguchi DoEs were used to identify, optimize, and refine buffer conditions for suppressing background. In the first exploratory DoE round, we observed that buffer conditions alone could modulate NSB across a wide response range, from complete suppression with retained control line signal to elevated membrane background and excessive test line signal, Supplementary **Figure S8** and **Tables S5-6**. In the next round, we parsed the NSB-governing parameters. A 1:32 plasma dilution was selected for yielding a visible test line with minimal nitrocellulose background and preserved membrane flow, whereas higher dilutions did not produce detectable test line signal; we repeated the experiments at 1:16 plasma dilution to validate the findings, Supplementary **Table S7**. The nitrocellulose membranes, response graphs, and relative contribution of parameters are given in **Figure 4c** and Supplementary **Figure S9-10**, and quantified in Supplementary **Table S8**. At 1:32 plasma dilution, relative contribution of NSB on the test line was reduced by ionic strength (55.5%) and buffer type (28.2%, driven by Tris). At 1:16 plasma dilutions, ionic strength remained the primary contributor (52.1%), suggesting that salt concentrations could be adjusted in our experiments to reduce NSB on the test line, which is consistent with plasma’s high content of charged proteins.^23^ Occasional high nitrocellulose membrane background was attributed to hydrophobic IgG-membrane interactions for experiments with low surfactant concentrations. Tris reduced NSB on the test line more effectively than phosphate buffers at 1:32 dilutions. No single factor dominated signal response; rather, effect sizes were distributed across parameters, indicating multifactorial background suppression, and potential parameter interactions and interdependencies, warranting further investigation. Thus, a final refinement DoE investigated interactions by perturbing Tris, Tween 20, and NaCl concentrations, Supplementary **Figure S11** and **Tables S9-10**. In concordance with observations from the previous DoE, Tris concentration had high relative contribution to background suppression (65.3%). To evaluate the presence of confounding two-way interactions, we plotted the factors as a function of each other, Supplementary **Figure S12**. Conditional mean plots showed a Tris-dependent NaCl response in the Tris × NaCl concentration pair, indicating that these parameters are coupled and must be jointly optimized rather than treated as independent.

We confirmed that Tris and NaCl did not impact the surface tension (and capillary flow), which was governed by Tween 20 in a concentration-dependant manner, Supplementary **Table S11-12**. Fluidic performance was highly sensitive to Tween 20 levels, with limited tolerance for its addition to preserve fluidic functionality. Consequently, DoE-guided optimization provided options for NSB reduction that relied on the addition of salts while keeping Tween 20 concentrations low (i.e., 0.01% v/v), Supplementary **Table S13**.

### Synchronizing capillaric MVT readout with each respective clinical threshold

DoE experiments so far identified two individual parameters that govern assay signal strength and analytical sensitivity (i.e., capture probe density and sample volume), and two collective parameters that govern background suppression (i.e., buffer composition and ionic strength). Armed with this knowledge, we empirically optimized the capillaric MVT readout by synchronizing with the clinical positivity thresholds.

To date, parameter identification, quantification, and separation was done in buffer and pre-pandemic pooled plasma as a negative control. In the absence of standardized positive controls for raAbs, we tested and optimized the capillaric MVTs with a small cohort of patient samples as the positive controls. We selected 6 patients with varying positivity (P1-6) along with the same pre-pandemic pooled plasma negative for all three antibodies (N1) based on their microplate ELISA results, **Figure 4d**. Microplate ELISA was also used to identify whether the impact of pre-antigen-bound raAbs in plasma would lead to potential false negatives, but a consistently higher signal for free raAbs in these patient samples ensured that the capillaric MVT immunoassay format did not need modification, Supplementary **Figure S13**.

With the DoE-selected parameters, we first used the collective assay parameters to reduce the background across the test strip, then used the individual capture probe densities to independently tune each assay’s visual readout to match the clinical thresholds, which was achieved within 5 empirical rounds, Supplementary **Figure S14** for full details, and **Figure 4e** for main findings. Anti-IFN-α and anti-IFN-ω assays were optimized to simultaneously meet their clinical thresholds and suppress visual NSB at the test line, whereas the anti-SCoV2 assay only required NSB suppression because its purpose was antibody detection rather than threshold-based classification. High Tris/NaCl initially caused false positives (round 1); increasing Tris suppressed NSB across all three assays but also eliminated true signals, resulting in false negatives (rounds 2-3). Finally, rebalancing Tris and NaCl concentrations restored the test line signals, and antigen spotting density restored specific signals for each assay at their clinical thresholds (rounds 4-5). The final assay conditions yielded antibody-specific signal intensities that matched the expected high-medium-low microplate ELISA rankings and enabled clinically relevant visual readout tunable using only a few experimental levers.

### Benchmarking and clinical assessment of capillaric MVTs with microplate ELISA

Capillaric MVTs were tested with a larger cohort of 41 patient plasma samples comprising of 5 samples positive for anti-IFN-ω raAbs, 6 positive for anti-IFN-ω raAbs, 3 positive for both anti-IFN-α and -ω raAbs, and 36 positive for anti-SCoV2 antibodies, Supplementary **Table S14**, to validate multiplexed visual readout for risk stratification.

The samples were blinded to the operator, and the yield of the chips was monitored by checking for the presence of a positive control line; instances where fabrication issues (poor printing and sealing) or poor chip-to-assay connections that resulted in a faint or lost control line were excluded and repeated. Then, according to the visual readout of the capillaric MVT, patients were stratified based on the presence/absence of high concentration anti-IFN raAbs.

The capillaric MVT readouts of all the patient plasma samples are given in Supplementary **Figure S15**, representative readouts showing visual thresholding of the individual targets are given in **Figure 5a**, with quantitative test line intensities in **Figure 5b**. Based on our criteria for control line visibility, only 3 capillaric MVTs were repeated when assembled and operated by a trained user, Supplementary **Figure S16**, demonstrating reliable fluidic performance when running patient samples. Compared to microplate ELISA as the clinical standard, the anti-IFN-α assay on the capillaric MVT correctly stratified all the patients except for two false negatives and a receiver operating characteristic (ROC)-area under the curve (AUC) of 0.97, followed by the anti-IFN-ω assay with no false readouts and an ROC-AUC of 1.00, and finally the anti-SCoV2 assay with one false positive and an ROC-AUC of 0.98, **Figure 5c**. Microplate ELISA optical density measured at 450 nm was compared to the test line intensities of the capillaric MVTs and a Pearson’s correlation of 0.93, 0.93, and 0.89 for the anti-IFN-α, anti-IFN-ω, and anti-SCoV2 tests, respectively, showed quantitative intensity correlation with the clinical standard, **Figure 5d**. Altogether, the capillaric MVT supported visual readout for stratification of patient plasma with quantitative agreement upon digitization relative to microplate ELISA.

**Figure 5.**
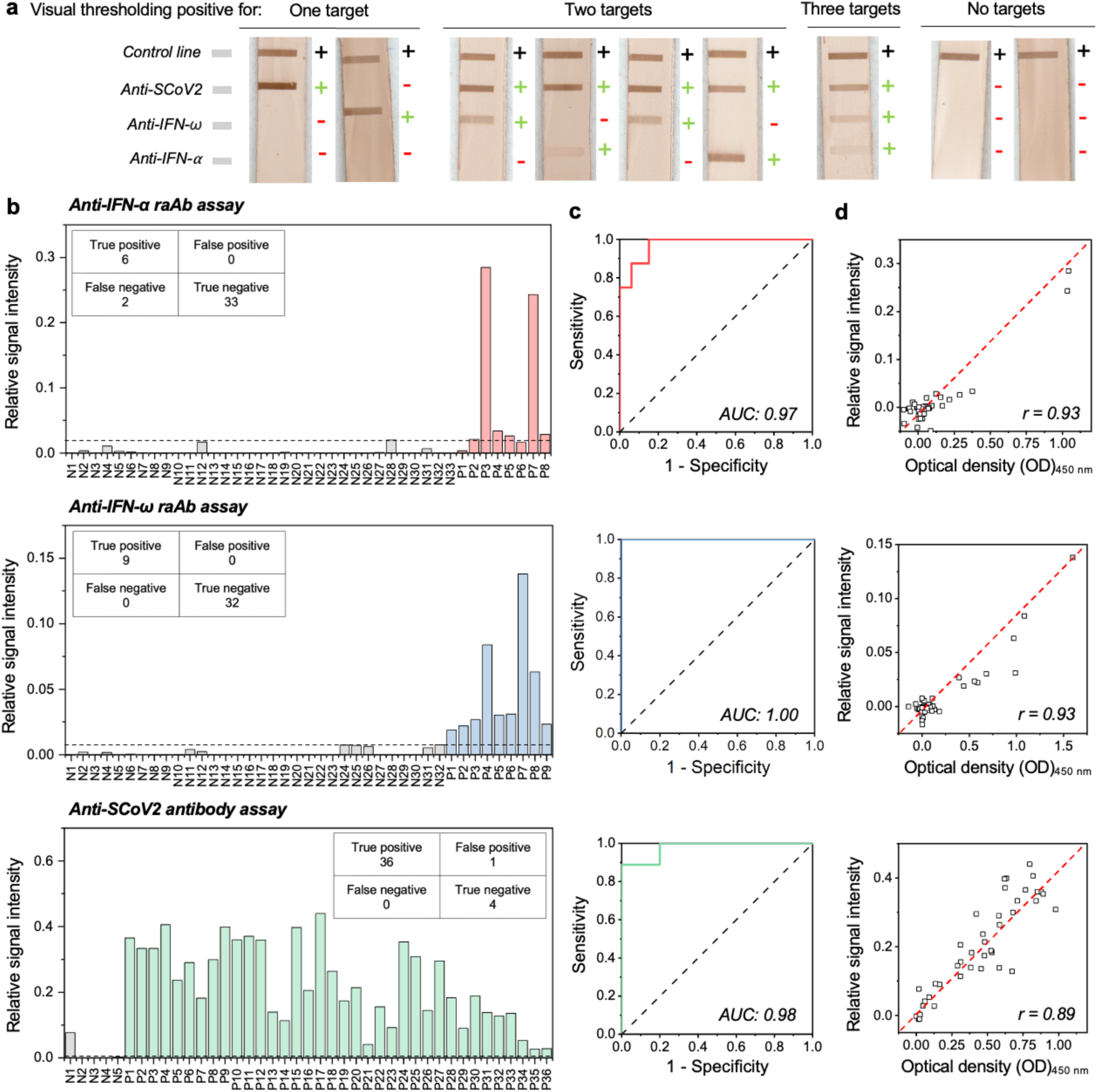
Benchmarking and clinical assessment of capillaric MVTs with microplate ELISA. (a) Representative nitrocellulose membranes from capillaric MVTs showing threshold-calibrated readouts across different targets used for patient stratification. Black + symbols show the presence of a control line, and coloured +/− symbols show the expected result of plasma samples based on microplate ELISA as the reference method. (b) Quantitative test line readouts of 41 clinical plasma samples for the detection of anti-IFN-α raAbs (top), anti-IFN-ω raAbs (middle), and anti-SCoV2 antibodies (bottom) in a triplex format. Dotted lines show the quantitative threshold between positive (P) and negative (N) samples given by the highest negative sample readout. Insets show a confusion matrix of the capillaric MVT readouts compared to microplate ELISA as the clinical standard. (c) Receiver operating characteristic (ROC) analysis compared with microplate ELISA and the corresponding area under the curve (AUC). (d) Comparison between capillaric MVT test line color intensity and microplate ELISA optical density revealing an overall strong correlation; inset shows the Pearson correlation (*r*) for the three assays, and the dotted red line shows a line of best fit.

## Discussion

We introduce a conceptual and experimental framework for the design and use of capillaric MVTs with readout synchronized to each respective clinical threshold for multiple, cross-reacting targets simultaneously, and we illustrate it for the triplex detection of anti-IFN-α and -ω raAbs, and anti-SCoV2 antibodies in patient plasma. Among 16 parameters considered in this study using statistical DoE, we first sought and identified individual parameters governing signal strength and analytical sensitivity, followed by collective parameters suppressing background across all three test lines. Using mouse and human antibodies in an off-chip array-format, we found that (i) the density of spotted capture antigen determined the visual threshold and linear range, and (ii) buffer composition and ionic strength could effectively suppress the background of all assays. Armed with this understanding, we used a high ionic strength buffer to minimize the background and adjusted the spotting density for each antigen to synchronize visual and clinical thresholds on the capillaric MVTs. We validated patient stratification with visual readout in a 41-patient study, achieving good concordance with microplate ELISA as the reference method while preserving correlated quantitative readouts.

Capture probe density governed signal strength and visual threshold for all assays, **Figure 2e**, in concordance with a capture probe-limiting regime.^8,9^ However, cross-reactivity between assays can alter binding signal intensities,^24^ as observed between anti-IFN-α antibodies and IFN-ω test lines; this interdependence was mitigated by constraining capture probe densities to ranges where visible cross-reactivity was negligible for MVTs, Supplementary **Figure S4**. The concentrations of raAb can be very high (ng-µg mL^−1^) and must be measured in a context of clinical thresholds that are comparatively elevated (ng mL^−1^). High concentration analytes can be measured with low capture probe densities and thus visual thresholds easily synchronize with clinical cutoffs while simultaneously reducing susceptibility to cross-reactivity. We reduced the capture probe density of IFN-ω from 56 to 14 ng mm^−2^ (i.e., 4-fold reduction, while an 8-fold reduction resulted in a ∼1000-fold decrease in visual threshold, see **Figure 2e**), thus synchronizing the visual and clinical thresholds under moderate sample dilution (i.e., 1:16 for patient plasma). The simultaneous reduction of visual threshold and cross-reactivity, shown here for raAbs, is expected to be generalizable to other high concentration targets that may also cross-react.

Background signal due to NSB on any assay test line are detrimental to MVTs because they create false positives. Interestingly, we observed that NSB on the test line was insensitive to capture probe density and sample volume in our experiments, but was instead influenced by plasma dilution, buffer composition, and ionic strength, and most notably by Tris and NaCl concentrations, **Figure 4c**. The optimized buffer in our study had high ionic strength (300 mM NaCl and 500 mM Tris), consistent with salt-dependent NSB expected from the abundance of charged proteins in plasma (e.g., complement proteins, heterophilic antibodies).^23,39,41^ Tris was more effective at reducing NSB, possibly due to weak competition for hydrogen bonding sites^42,43^ in raAb tests. While our findings indicate that raAb detection in plasma benefits from high ionic strength buffers, extending our approach to other assays, such as antigen detection, will require reidentifying collective parameters that govern NSB for the new targets.

Among the 41 patient plasma samples used to benchmark the capillaric MVTs, 14 tested positive for high levels of either one or both anti-IFN raAbs, **Figure 5a-b**, Supplementary **Figure S15**. Of these, three patients developed severe or critical COVID-19, Supplementary **Table S14**, consistent with the established association between neutralizing anti-type 1 IFN raAbs and increased risk of severe disease.^12–14^ Although our study retrospectively evaluated banked clinical samples, future prospective studies in larger, independent cohorts are needed to further validate assay performance and determine how capillaric MVTs can be integrated into clinical triage workflows for risk stratification and treatment decision-making.

Taken together, the results support the feasibility of threshold-calibrated MVTs. The proposed conceptual and experimental framework for their design can be described as follows:

(1) Use DoE to identify universal buffer that minimize background and NSB without interfering with the assay signal for all targets;
(2) Use DoE to identify the parameters that individually control the assay signal strength (capture probe density being the primary candidate parameter);
(3) Optimize buffer composition to minimize background and NSB for each target;
(4) Tune assay signal individually to match the clinical threshold for each target.

And the conditions that the assays must meet individually and collectively to enable MVTs are:

a. The LOD of each assay is markedly lower than the clinical threshold so that it can be tuned to meet the threshold;
b. The targets are mutually compatible in a shared buffer system that effectively mitigates background and NSB for all targets under consideration;
c. The reagents are compatible and visible cross-reactivity is much lower than specific binding;
d. There is an independent parameter for each target that can be tuned to synchronized visual threshold to the clinical cutoff.

By following this framework, the rational design of clinically relevant threshold-calibrated MVTs can be completed for capillaric assays, as well as for other assay formats with visual readout, including LFAs. As assay signal is commonly governed by capture probe density, it is expected to be available as a parameter to tune the visual threshold individually. Hence the condition that predictably will be the most challenging to meet is a buffer formulation that suppresses the background for all the selected targets, or conversely, to identify targets that satisfy the “low background criteria” for a given buffer formulation. Future work is needed to determine which type of targets, and which specific ones, are mutually compatible for tandem measurements. In the case of LFAs, additional constraints such as reagent drying and absence of wash buffers must be met, which could further constrain the operating window for LFA-based MVTs. The ELISA-chip and multi-step assays in general benefit from greater freedom of design and flexibility, including rinsing steps, and could for instance consider epitope depletion^44^ to convert a high concentration target into a low concentration one facilitate its inclusion on an MVT with low concentration targets.

Visually interpretable self-contained MVTs are particularly well-suited to reflex testing workflows, in which rapid, low-cost, point-of-care screening identifies high-risk patients for quantitative confirmation testing. In this setting, assays can prioritize analytical sensitivity to minimize false negatives while tolerating a number of false positives, as these can be resolved through subsequent testing, also at the point-of-care, or in a central lab. Indeed, while MVTs are intrinsically visual yes/no tests, they are also compatible with quantitative image-based analysis, and dual-readout workflows where for instance, only ambiguous or positive tests are selected for quantification, could help improve overall testing accuracy while minimizing the impact of lighting conditions, surface texture, and user variability.^45–47^ Workflows with reflex testing can increase capacity for screening while still enable timely therapeutic intervention upon confirmation for acute diseases such as COVID-19 (e.g., administration of Paxlovid within the first 1-3 days of infection), all while strengthening future pandemic preparedness.^48,49^

Single-use capillaric ELISA-chip format enables decentralized testing without batch processing or digital recording, which may also benefit privacy-sensitive settings. Capillaric MVTs are already compatible with low-precision pipettes,^26^ 3D-printed with low-cost (US$150-600) LCD printers for global health users,^28^ and future work towards on-chip reagent storage could further simplify operation for untrained users.^50^ Beyond anti-IFN raAbs, MVTs could be broadly applicable for other applications requiring urgent risk stratification, including septic shock (procalcitonin, preseptin, and C-reactive protein),^51,52^ myocardial infarction (troponin I, pro-brain natriuretic peptide, and cardiac troponin T),^53–55^ or multi-antigen serotests for tuberculosis,^56^ as well as non-urgent conditions such as autoimmune diseases (e.g., rheumatoid arthritis, targets including anti-cyclic citrullinated peptide, rheumatoid factor).^57^ MVTs calibrated to clinical thresholds could be used as stand-alone, and used for triage for quantification with a reader or as independent tests, highlighting their broad potential for improving patient care thanks to point-of-care testing and more efficient clinical workflows.

## Materials and Methods

### Materials

#### Immunoassay reagents

##### Capillaric MVTs

Human IFN-α 2a carrier-free protein (Cat. #11101-2, lot #7346, PBL Assay Science, Piscataway, New Jersey, United States), anti-human IFN-α clone MMHA-2 mouse monoclonal antibody (Cat. #21100-2, lot #7291, PBL Assay Science, Piscataway, New Jersey, United States), recombinant human IFN-ω protein (Cat. #NBP2-35893, lot #m10609112, R&D Systems, Minneapolis, Minnesota, United States), anti-human IFN-ω mouse monoclonal antibody (Cat. #NBP3-06154, lot #HB09N03003-B, R&D Systems, Minneapolis, Minnesota, United States), SARS-CoV-2 spike protein S1+S2 (Cat #40589-V08B, lot #LC14MA1306, Sino Biological, Inc., Beijing, China), SARS-CoV-2 spike S1 subunit mouse monoclonal antibody (Cat. #MAB105403, lot #CNFM0121031, R&D Systems, Minneapolis, Minnesota, United States), goat anti-mouse IgG biotinylated antibody (Cat. #BAF007, lot #WYQ0721071, R&D Systems, Minneapolis, Minnesota, United States), preadsorbed biotinylated goat anti-human IgG Fc (Cat. #AB98618, lot #1017182-2, Abcam, Cambridge, United Kingdom), Pierce streptavidin poly-horseradish peroxidase (pHRP) (Cat. #21140, lot #XJ360080, ThermoFisher Scientific, Waltham, Massachusetts, United States), SIGMAFAST 3,3’-diaminobenzidine (DAB) tablets (Cat. #D4293, lot #SLCG5357, Sigma-Aldrich, Oakville, Ontario, Canada), bovine serum albumin (BSA) (Cat. #001-000-162, lot #162191, Jackson ImmunoResearch Labs, West Grove, Pennsylvania, United States), BSA-biotin (Cat. #A8549, Sigma-Aldrich, Oakville, Ontario, Canada), citrated normal human pooled plasma (Cat. #PL080322MU, lot #BC80322PHP, BioChemed Services, Winchester, Virginia, United States), citrated defibrinated human pooled plasma (Cat. #P9523, lot #SLBX8880, Sigma-Aldrich, Oakville, Ontario, Canada), Tween 20 (Cat. #P7949, lot #SLBX0835, Sigma-Aldrich, Oakville, Ontario, Canada).

##### Microplate ELISA

Human IFN-α protein (Cat. #11105-1, lot #7361, PBL Assay Science, Piscataway, New Jersey, United States), recombinant human IFN-ω protein (Cat. #11395-1, lot #7486, PBL Assay Science, Piscataway, New Jersey, United States), SARS-CoV-2 spike protein S1+S2 (Cat. #40589-V08B, lot #LC14MA1306, Sino Biological, Inc., Beijing, China), goat anti-human IgG (H+L) secondary antibody with HRP conjugate (Cat. #A18805, lot #77-66-071020, ThermoFisher Scientific, Waltham, Massachusetts, United States), BLOTTO non-fat powdered milk (Cat. #B501, Rockland Immunochemicals, Pottstown, Pennsylvania, United States), 3,3’,5,5’-tetramethylbenzidine (TMB) substrate (Cat. #T0440, lot #0000330433, Sigma-Aldrich, Oakville, Ontario, Canada), sulfuric acid (Cat. #351293-500, lot #115107, Fisher Scientific, Saint-Laurent, Quebec, Canada).

#### Capillaric MVT 3D printing and assembly

Poly(ethylene glycol) diacrylate (PEGDA)-250 (Cat. #475629, lot #MKCS0146, Sigma-Aldrich, Oakville, Ontario, Canada), diphenyl(2,4,6-trimethylbenzoyl)phosphine oxide (TPO) (Cat. #415952, lot #MKCK2346, Sigma-Aldrich, Oakville, Ontario, Canada), 2-isopropylthioxanthone (ITX) (Cat. #I067825G, lot #ZNNQE-KT, TCI America, Portland, Oregon, United States), pentaerythritol tetraacrylate (PETTA) (Cat. #408263, lot #MKCR5556, Sigma-Aldrich, Oakville, Ontario, Canada), acrylic acid (Cat. #147230, lot #SHBD0923V, Sigma-Aldrich, Oakville, Ontario, Canada), isopropyl alcohol (IPA) (Cat. #RPD99, Rapid Protectant, Concord, Ontario, Canada), nitrocellulose membranes (Cat. #VIV1202503R, lot#1205178320, Pall Corporation, New York, United States), glass fiber conjugate pad (Cat. #GFCP103000, lot #778051, Sigma-Aldrich, Oakville, Ontario, Canada), absorbent pads grade 320 (Cat. #3208-2020, lot #115248, Ahlstrom-Munksjo Chromatography, Lab Filtration Papers, Connecticut, United States), microfluidic sealing tape (Cat. #9795R, 3M, Perth, Ontario, Canada).

#### Additional reagents

Sodium phosphate dibasic heptahydrate (Cat. #S-373B, lot #874580, Fisher Scientific, Saint-Laurent, Quebec, Canada), sodium phosphate monobasic monohydrate (Cat. #S369-500, lot #072399, Fisher Scientific, Saint-Laurent, Quebec, Canada), Triton X-100 (Cat. #BP151-500, lot #051126, Fisher Scientific, Saint-Laurent, Quebec, Canada), Surfact-Amps NP-40 (Cat. #28324, lot #MF159823, ThermoFisher Scientific, Waltham, Massachusetts, United States), Brij-35 (Cat. #A15809, lot #10162641, Alfa Aesar, Haverhill, Massachusetts, United States).

All assay reagents were prepared using 1X phosphate-buffered saline (PBS) (pH ∼7.4) supplemented with Tween 20 and BSA unless otherwise specified. All other solutions were prepared using water from a MilliQ system (resistivity: 18 MΩ cm; Millipore).

### Taguchi design of experiments and statistical analysis

Experiments were designed with the JMP Statistical Discovery (v17.2, SAS Institute, North Carolina, United States) software using the built-in Taguchi arrays or the augmented design module for custom arrays. Orthogonality and (multi)collinearity of the proposed designs were evaluated to consider the impact of correlated variables in the designed experiment. Results were analyzed in JMP using analysis of variance (ANOVA) to quantify the relative contribution and effect size of parameters and their interactions. Data was plotted to show the mean of a given effect and error bars represent the standard error across grouped experiments; details are given in Supplementary **Note S1**.

### Nitrocellulose membrane assay preparation

The assay was based on lateral flow nitrocellulose membranes and was prepared as described previously.^25–28^ Briefly, nitrocellulose membranes were cut to 3 × 12 mm^2^ (width × length) with a pointed tip using a film cutter (Cameo 3, Silhouette Portrait, Lindon, United States) and inkjet spotted (sciFLEXARRAYER SX, Scienion) with 2.5 × 1 mm^2^ (width × length) test and control lines spaced 2 mm apart center-to-center. For the test line, protein solutions were prepared in 0.22 µm filtered 1X PBS buffer and loaded into the piezo-driven capillary and then programmed to release 350 pL droplets in a 25 × 4 (width × length) line array over several passes (optimized using DoE); each pass spotted alternating positions on the line array so that droplets could dry between passes. Similarly, 50 µg mL^−1^ of BSA-biotin was spotted with 8 passes in alternating positions of the line array for the control line. The spotted membranes were dried at 37°C for 1 h, then blocked by dipping and then submerging the membranes in a tray containing ∼5 mL blocking buffer (composition optimized using DoE) and then placed on an orbital shaker at 100 rpm for 1 h at room temperature. The membranes were dried again for 1 h at 37°C, then stored with desiccant in a Parafilm M sealed container at 4°C overnight and used within 3 days of protein spotting.

### Off-chip array-format DoE

The off-chip nitrocellulose assay was designed by sandwiching strips of the spotted membrane between absorbent pads held together using a binder clip, then dipped into the wells of a conventional flat-bottom 96-well plate pre-loaded with assay reagents to encode the DoE experiments. Rows of wells were loaded with assay reagents at defined concentrations and volumes, interspersed with wash steps, and nitrocellulose test strips were manually transferred between wells after each reagent flowed over the membrane by capillary flow.

### Ink preparation, 3D printing, and assembly of capillaric MVTs

Capillaric MVTs were designed in AutoCAD (Autodesk 2025) and exported as .stl files for slicing in a third-party software, CHITUBOX, at a layer thickness of 20 µm with the following print settings at an irradiance of 2.23 mW cm^−2^: 10 s base exposure time for 5 base layers; 1.8 s normal exposure time; 5 transition layers; 3 s rest time between ascent and descent of the build plate; 3 mm ascent distance between layers; 80 mm min^−1^ ascent speed; 210 mm min^−1^ descent speed, resulting in an estimated print time of ∼35 min for capillaric MVTs. The devices were 3D-printed using a custom poly(ethylene glycol) diacrylate (PEGDA)-based ink, denoted as PLInk, as described previously.^28^ Briefly, the ink was prepared by adding 0.5% (wt/wt) TPO photoinitiator, 1.5% (wt/wt) ITX photoabsorber, and 2% (wt/wt) PETTA crosslinker to PEGDA-250 and mixed under magnetic stir for 30 min at room temperature or until fully dissolved, then stored in glass amber bottles and kept at room temperature until use. Capillaric MVTs were 3D-printed using the Elegoo Mars 4 Ultra 9K (ELEGOO, Shenzhen, China) LCD 3D printer equipped with a 405 nm illumination wavelength and featuring 8520 × 4320 @ 36.8 M pixels that are 18 × 18 µm^2^ in size. Printed devices were washed with IPA and dried under a stream of compressed nitrogen gas. Then, the capillaric MVTs were submerged in an IPA solution containing 10 mM acrylic acid and 10 mM TPO photoinitiator, and post-cured in a UV lamp chamber (CureZone, Creative CADWorks, Concord, Canada) for 30 s, then dried again under a stream of compressed nitrogen gas and sealed to encapsulate the microchannels using a pressure adhesive microfluidic sealing tape. The devices were assembled by sandwiching the spotted nitrocellulose assay membranes between absorbent pads laser cut to be 10 × 2.4 mm^2^ (width × length), mounted onto the capillaric MVTs using an in-place 3D-printed guide, and the top drainage channel was also mounted with a 1.5 × 3.5 mm^2^ (width × length) glass fiber, then clamped in place using a 1-inch binder clip. Devices were either loaded and run immediately with assay reagents or stored in a vacuum-sealed bag containing an oxygen absorber until use. Assay reagents were prepared in buffer solutions (optimized using DoE), and the assay substrate solution was prepared by dissolving SIGMAFAST^TM^ DAB (3,3’-diaminobenzidine) tablets in 5 mL MilliQ water and 0.22 µm filtered before use.

### Dynamic tensiometry

The surface tension of buffer solutions was measured using a dynamic tensiometer (DCAT11, DataPhysics Instruments GmbH). Briefly, 50 mL of buffer solutions was poured into a glass crystallizing dish and mounted onto the calibrated instrument balance. A platinum-iridium plate (PT 11, length: 10 mm, width: 19.9 mm, thickness: 0.2 mm, part no. 2000323, DataPhysics Instruments GmbH) was used for surface and interfacial tension measurements according to the Wilhelmy plate method as it descended and rose through the liquid interface, then recorded the average surface tension over the plate’s ascent time until the values were within 0.03 mN m^−1^ of each other. Between measurements, the plate was cleaned by rinsing with MilliQ water and 95% ethanol, followed by blow torching until the entire plate turned red and then cooling in a stream of compressed air to ensure the plate was clean and not wetted between measurements. All measurements were performed at 22-23°C.

### Microplate ELISA protocol

Microplate ELISAs were performed by coating 384-well MaxiSorp plates with 100 µL of either 10 ng mL^−1^ recombinant IFN-α, 1 ng mL^−1^ recombinant IFN-ω, or 250 ng mL^−1^ SARS-CoV-2 S1 subunit spike protein solutions prepared in a 50 mM carbonate buffer (pH 9.6) and incubated overnight at 4°C. Then, the wells were washed with PBST 0.1%, followed by 2 h blocking at 37°C with the wash buffer supplemented with 5% non-fat milk powder. The wells were washed again, then incubated with 1:100 dilutions of patient plasma and controls for 1 h at room temperature. Samples and controls were loaded in triplicate on both antigen-coated and uncoated wells. The wells were washed again. Goat anti-human IgG conjugated with HRP was prepared to a 750 ng mL^−1^ solution in PBST 0.1% supplemented with 3% BSA and 100 µL was added to each well followed by 1 h incubation at room temperature and without exposure to light. Finally, the plates were washed, and 50 µL of TMB substrate was added. After 5 min, or until the blanks began to develop colour, 50 µL of 0.16 M sulphuric acid was added as the stop solution. The optical density was read using a NanoDrop UV-Vis spectrophotometer (NanoDrop@ND-1000, NanoDrop Technologies, Wilmington, Delaware, United States) at 450 nm with a 1 mm path length.

### Collection and processing of clinical samples

Human plasma from individual donors was collected and processed at the McGill University Health Center (MUHC) in accordance and with approval granted by the MUHC Research Ethics Board (MP-37-2021-7489) from April 2021 to April 2022. Participants were informed and consented to being involved in the study and provided potentially identifying information; all samples were deidentified from their individual donors prior to testing and analysis. Samples were collected as whole blood in sodium heparin or citrate anticoagulant tubes, then centrifuged at 850 g for 10 min at room temperature. The plasma layer was collected, aliquoted into 250 or 500 µL volumes, and then stored in −80°C until use.

### Videos and image processing

Videos of the capillaric MVTs loaded and run with dyed buffer solutions were recorded to characterize the reagent flow using a Sony α7R III camera with a macro lens (Sony FE 90 mm F2.8 Macro G OSS Lens). Nitrocellulose membranes were imaged using a flatbed desktop scanner (Epson Perfection V600) using the Epson Scan software (v3.9.3.1, Seiko Epson Corporation, Nagano, Japan) at 1200 dpi in 48-bit RBG format, then imported to ImageJ2 for measuring 16-bit grayscale colorimetric line intensity ranging from 0-65,535 gray values. The readouts were first adjusted to flip the colour scale to render “true white” as 0 and “true black” as 65535, then rescaled between 0-1 relative signal intensities. Signal intensities are given by the colorimetric intensity of the test line subtracted by the average intensity measured 1 mm above and below to account for the local background. For the binding curves, signal intensities at different analyte concentrations are fitted to a 4-point logistic curve, and the LOD is given as the concentration point on the fitted curve where the signal intensity is the sum of the mean of the blank plus three standard deviations.

## Acknowledgements

This work was supported by the Natural Sciences and Engineering Research Council of Canada (NSERC) Discovery Grant (RGPIN-2022-05171) and the McGill University MI4 Seed Fund Grant. H.S. acknowledges support from the Canadian Institutes of Health Research (CIHR) Canada Graduate Research Scholarship – Master’s program (CGS-M), the Fonds de recherche du Québec – Nature et technologies (FRQNT) master’s research scholarship (no. 334837), and the NSERC Canada Graduate Research Scholarship – Doctoral program (CGS-D). D.C.V. acknowledges support from the Fonds de Recherche du Québec – Santé (FRQS) (no. 251575). Operational support for patient sample collection was received from the Public Health Agency of Canada: COVID-19 Immunity Task Force, the Montreal General Hospital Foundation, and the FRQS Biobanque québécoise de la COVID-19 (BQC19). D.J. acknowledges support from a Canada Research Chair in Bioengineering (CRC-232159).

## Author contributions

H.S. designed and optimized the nitrocellulose assays and capillaric MVTs. H.S. and S.B. optimized and performed the microplate ELISAs. S.B., L.C., and D.C.V. collected, processed, and banked clinical data and patient plasma samples. H.S., A.N., and D.J. planned the experiments and interpreted the data. H. S. prepared the figures and analyzed the results. H.S., A.N., and D.J. wrote the manuscript with feedback from all co-authors. D.C.V. and D.J. provided funding, and D.J. conceptualized, supervised, and administered the project.

## Data availability

Data not presented in the article or supplementary material will be available upon request.

