## Supplementary Information for "Multiplexed visual test with readout synchronized to each respective clinical threshold for rogue anti-cytokine autoantibodies"

**Table of Contents**

1. Brief background on DoE calculations 3
   1.1. Summary of quantifying parameters, their interactions, and their relative contribution to the assay signal in DoE 3
2. Identification of key individual parameters governing assay performance and assay interdependency 4
   2.1. Reagent concentration and volume screening and quantification of parameter weights 4
   2.2. Screening of wash steps and quantification of parameter weights 9
   2.3. Cross-reactivity in MVTs 11
3. 3D-printed capillaric MVTs 12
   3.1. Logistic fit model, limit of detection, and visual threshold 12
4. Identification of key collective parameters governing background suppression 13
   4.1. Potential factors contributing to NSB 13
   4.2. Diluent buffer composition to supress visual background and quantification of parameter and interaction weights 15
   4.3. Surface tension of buffer components 26

5. Benchmarking and clinical assessment of capillaric MVTs 28
5.1. Patient cohort information 28
5.2. Pre-antigen-bound raAbs in circulation 31
5.3. Empirical rounds to synchronize visual and clinical thresholds on capillaric MVTs 32
5.4. Capillaric MVT readouts of clinical samples 33
References 34

### 1. Brief background on DoE calculations

#### *1.1. Summary of quantifying parameters, their interactions, and their relative contribution to the assay signal in DoE*

Parameters and their interactions were quantified with ANOVA, effect sizing, and relative contribution. If a given parameter accounted for a substantial portion of the variance in the assay response, then it had a highly weighted contribution to the overall assay response. These quantifications were only valid if relationships between parameters are orthogonal, verified using orthogonality and collinearity tests in JMP (e.g., variance inflation factor).

We first computed the total sum of squares (SS) and ANOVA, then we used SS to obtain the effect size and relative contribution for each parameter and interaction being investigated, as follows:

$$SS = \sum_{i=1}^{n} (y_i - \bar{y})^2$$

Where $y_i$ is the value in sample $i$ over $n$ total observations, and $\bar{y}$ is the mean value of that sample.

The effect size ($\eta^2$) explains the proportion of the *total* variance (considering all factors and error) by a given factor, making it generalizable across experiments, and is computed as follows:

$$\eta^2 = \frac{SS_{factor}}{SS_{total}} = \frac{SS_{factor}}{\sum SS_{all\,factors} + SS_{error}}$$

Relative contribution explains the proportion of *model* variance (not considering the error) by a given factor, making it useful to rank factors in each experiment, and is computed as follows:

$$\text{Relative contribution (\%)} = \frac{SS_{factor}}{\sum SS_{all\,factors}} \times 100\%$$

We did not perform post-hoc tests to assess the significance of factors as *p*-values are not insensitive to sample size, which was variable across different DoE experiments and was small due to the few number of experiments performed in DoE, resulting in unstable *p*-values as effects were added/dropped from the model; meanwhile, relative contribution and effect sizes remained conserved.

#### 2. Identification of key individual parameters governing assay performance and assay interdependency

##### *2.1. Reagent concentration and volume screening and quantification of parameter weights*

To identify parameters that drove assay performance, including visual threshold, LOD, and linear range, we first evaluated reagent concentrations and volumes, the latter encoding incubation times on a CC as the reagent flows at a given flow rate over the nitrocellulose membrane. For a standard immunoassay, this included the capture probe density, dAb concentration and volume, enzyme concentration and volume, and substrate volume, summing up to 7 continuous factors. Among the available DoE Taguchi orthogonal array designs,[1] we selected the Taguchi $L_{27}(3^{13})$ to perturb up to 13 continuous factors at three levels in 27 experiments. Smaller orthogonal arrays had either fewer levels or factors. 27 experiments meant 26 degrees of freedom: 7 were assigned to continuous parameters and the remaining 19 were used to evaluate potential confounding 2-way interactions and the error. Parameter ranges were selected based on prior knowledge of ELISA-chips and microplate ELISAs. The orthogonal array experimental configuration is given in **Table S1**, nitrocellulose membranes and quantitative readouts in **Figure S1**, quantified parameter weights for the array repeated at two sample concentrations (5 and 500 ng $mL^{-1}$) in **Table S2**, and the DoE response graphs for each weighted parameter in **Figure S2**.

**Table S1. Reagent concentration and volume optimization using Taguchi $L_{27}(3^{13})$ orthogonal array experimental configuration***

| Exp# | Capture probe density (ng mm$^{-2}$) | dAb concentration (µg mL$^{-1}$) | Enzyme concentration (µg mL$^{-1}$) | Sample volume (µL) | dAb volume (µL) | Enzyme volume (µL) | Substrate volume (µL) |
|---|---|---|---|---|---|---|---|
| **1** | 1.4 | 0.2 | 0.2 | 25 | 25 | 25 | 25 |
| **2** | 1.4 | 0.2 | 0.2 | 25 | 75 | 75 | 75 |
| **3** | 1.4 | 0.2 | 0.2 | 25 | 225 | 225 | 225 |
| **4** | 1.4 | 1 | 1 | 75 | 25 | 25 | 25 |
| **5** | 1.4 | 1 | 1 | 75 | 75 | 75 | 75 |
| **6** | 1.4 | 1 | 1 | 75 | 225 | 225 | 225 |
| **7** | 1.4 | 5 | 5 | 225 | 25 | 25 | 25 |
| **8** | 1.4 | 5 | 5 | 225 | 75 | 75 | 75 |
| **9** | 1.4 | 5 | 5 | 225 | 225 | 225 | 225 |
| **10** | 2.8 | 0.2 | 1 | 225 | 25 | 75 | 225 |
| **11** | 2.8 | 0.2 | 1 | 225 | 75 | 225 | 25 |
| **12** | 2.8 | 0.2 | 1 | 225 | 225 | 25 | 75 |
| **13** | 2.8 | 1 | 5 | 25 | 25 | 75 | 225 |
| **14** | 2.8 | 1 | 5 | 25 | 75 | 225 | 25 |
| **15** | 2.8 | 1 | 5 | 25 | 225 | 25 | 75 |
| **16** | 2.8 | 5 | 0.2 | 75 | 25 | 75 | 225 |
| **17** | 2.8 | 5 | 0.2 | 75 | 75 | 225 | 25 |
| **18** | 2.8 | 5 | 0.2 | 75 | 225 | 25 | 75 |
| **19** | 5.6 | 0.2 | 5 | 75 | 25 | 225 | 75 |
| **20** | 5.6 | 0.2 | 5 | 75 | 75 | 25 | 225 |
| **21** | 5.6 | 0.2 | 5 | 75 | 225 | 75 | 25 |
| **22** | 5.6 | 1 | 0.2 | 225 | 25 | 225 | 75 |
| **23** | 5.6 | 1 | 0.2 | 225 | 75 | 25 | 225 |
| **24** | 5.6 | 1 | 0.2 | 225 | 225 | 75 | 25 |
| **25** | 5.6 | 5 | 1 | 25 | 25 | 225 | 75 |
| **26** | 5.6 | 5 | 1 | 25 | 75 | 25 | 225 |
| **27** | 5.6 | 5 | 1 | 25 | 225 | 75 | 25 |

*Columns 8-13 are empty and used for assessing parameter interactions, so while they do not encode an experimental condition, they are included in the design for extra degrees of freedom.

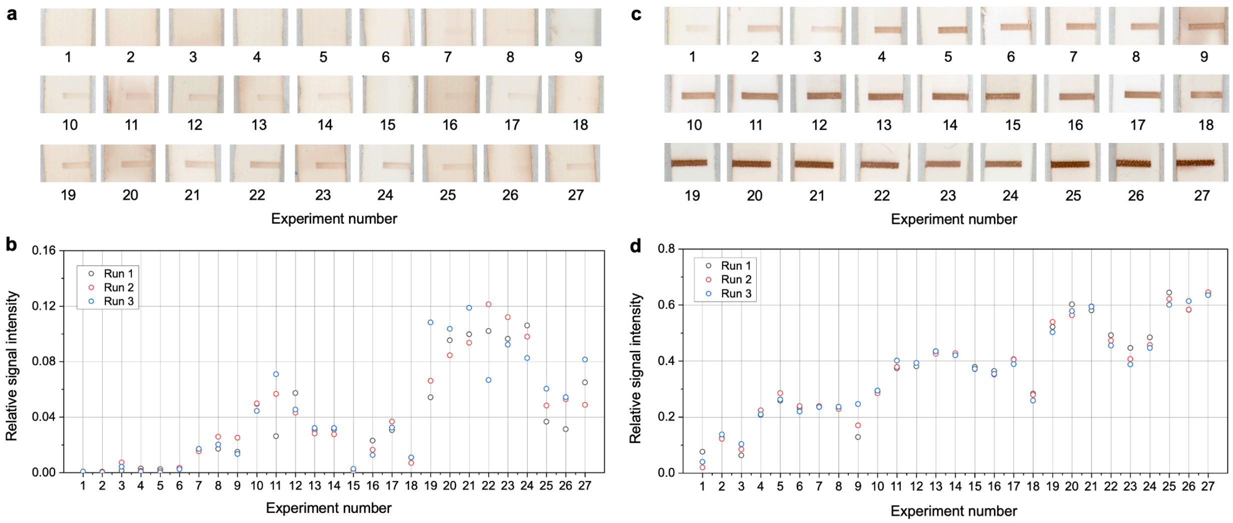

**Figure S1. Reagent concentration and volume Taguchi $L_{27}(3^{13})$ orthogonal array results.** (a, c) Nitrocellulose membrane test lines and (b, d) the corresponding relative signal intensity of the test line with different reagent concentrations and volumes using the Taguchi $L_{27}(3^{13})$ orthogonal array at both sample concentrations near the LOD (left), and in the linear range (right) for the mouse anti-human-IFN-α assay.

**Table S2. ANOVA, effect size, and percent relative contribution of assay parameters for reagent concentration and volume**

| Near the LOD, sample antibody concentration of 5 ng $mL^{-1}$ | | | | | |
|---|---|---|---|---|---|
| **Factor** | **Degree of freedom** | **Sum of squares** | **F ratio** | **$\eta^2$ effect size [0-1]** | **Relative contribution (%)** |
| **Capture probe density (ng $mm^{-2}$)** | 1 | 0.025 | 256.7 | 0.78 | 82.7 |
| **dAb concentration (µg $mL^{-1}$)** | 1 | 0.001 | 12.1 | 0.04 | 3.9 |
| **Enzyme concentration (µg $mL^{-1}$)** | 1 | 0.0002 | 1.9 | 0.006 | 0.6 |
| **Sample volume (µL)** | 1 | 0.004 | 39.2 | 0.12 | 12.6 |
| **dAb volume (µL)** | 1 | 2e-7 | 0.002 | 0 | 0.0007 |
| **Enzyme volume (µL)** | 1 | 4e-5 | 0.4 | 0.001 | 0.1 |
| **Substrate volume (µL)** | 1 | 7e-6 | 0.07 | 0.0002 | 0.02 |
| **In the linear range, sample antibody concentration of 500 ng $mL^{-1}$** | | | | | |
| **Factor** | **Degree of freedom** | **Sum of squares** | **F ratio** | **$\eta^2$ effect size [0-1]** | **Relative contribution (%)** |
| **Capture probe density (ng $mm^{-2}$)** | 1 | 0.570 | 107.2 | 0.79 | 92.4 |
| **dAb concentration (µg $mL^{-1}$)** | 1 | 0.013 | 2.4 | 0.02 | 2.1 |
| **Enzyme concentration (µg $mL^{-1}$)** | 1 | 0.024 | 4.4 | 0.03 | 3.8 |
| **Sample volume (µL)** | 1 | 0.006 | 1.2 | 0.009 | 1.0 |
| **dAb volume (µL)** | 1 | 7e-5 | 0.01 | 0.0001 | 0.01 |
| **Enzyme volume (µL)** | 1 | 0.001 | 0.2 | 0.001 | 0.2 |
| **Substrate volume (µL)** | 1 | 0.003 | 0.5 | 0.004 | 0.5 |

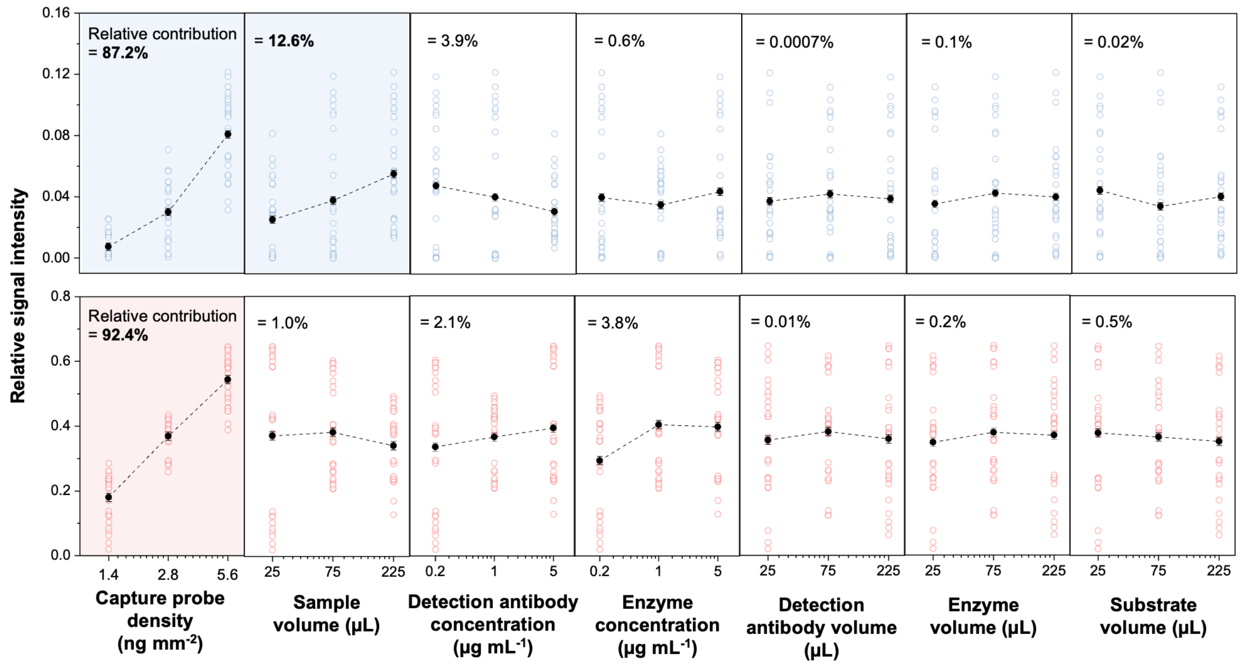

**Figure S2. Response graphs of reagent concentration and volume Taguchi $L_{27}(3^{13})$ orthogonal array.** Scatter plots illustrating the effect of each parameter on assay signal; insets show the corresponding quantified relative signal intensities. Responses for analyte concentrations both near the LOD (top, blue) and in the linear range (bottom, red) are shown for the mouse anti-human-IFN-α assay. Solid data points show the mean ± standard error across grouped runs; hollow data points show the results across all experimental runs. Relative contributions are shown inset.

##### *2.2. Screening of wash steps and quantification of parameter weights*

We evaluated whether wash steps had an impact on the signal intensities and determine whether certain washes needed to be prioritized or if some could be dropped to reduce the assay runtime. Four conventional wash steps in an ELISA were considered at high and low volume, and their effect quantified. The orthogonal array experimental configuration is given in **Table S3**, quantified parameter weights for the array in **Table S4**, and nitrocellulose membranes, quantitative readouts, and the DoE response graphs for each weighted parameter in **Figure S3**.

**Table S3. Screening of wash steps using Taguchi $L_8(2^4 \times 4^1)$ orthogonal array experimental configuration***

| Exp# | Wash 1 volume (µL); between sample and dAb | Wash 2 volume (µL); between dAb and enzyme | Wash 3 volume (µL); between enzyme and substrate | Wash 4 volume (µL); after substrate |
|---|---|---|---|---|
| **1** | 20 | 20 | 20 | 20 |
| **2** | 20 | 20 | 100 | 100 |
| **3** | 20 | 100 | 20 | 100 |
| **4** | 20 | 100 | 100 | 20 |
| **5** | 100 | 20 | 20 | 100 |
| **6** | 100 | 20 | 100 | 20 |
| **7** | 100 | 100 | 20 | 20 |
| **8** | 100 | 100 | 100 | 100 |

*Column 5 is empty and used for assessing parameter interactions, so while it does not encode an experimental condition, it is included in the design for extra degrees of freedom.

**Table S4. ANOVA, effect size, and percent relative contribution of assay parameters for screening of wash steps**

| Factor | Degree of freedom | Sum of squares | F ratio | $\eta^2$ effect size [0-1] | Relative contribution (%) |
|---|---|---|---|---|---|
| **Wash 1 volume; between sample and dAb (µL)** | 1 | 0.0001 | 0.3 | 0.02 | 2.7 |
| **Wash 2 volume; between dAb and enzyme (µL)** | 1 | 0.0008 | 1.8 | 0.14 | 18.8 |
| **Wash 3 volume; between enzyme and substrate (µL)** | 1 | 0.0032 | 7.4 | 0.59 | 77.7 |
| **Wash 4 volume; after substrate (µL)** | 1 | 3e-5 | 0.07 | 0.006 | 0.7 |

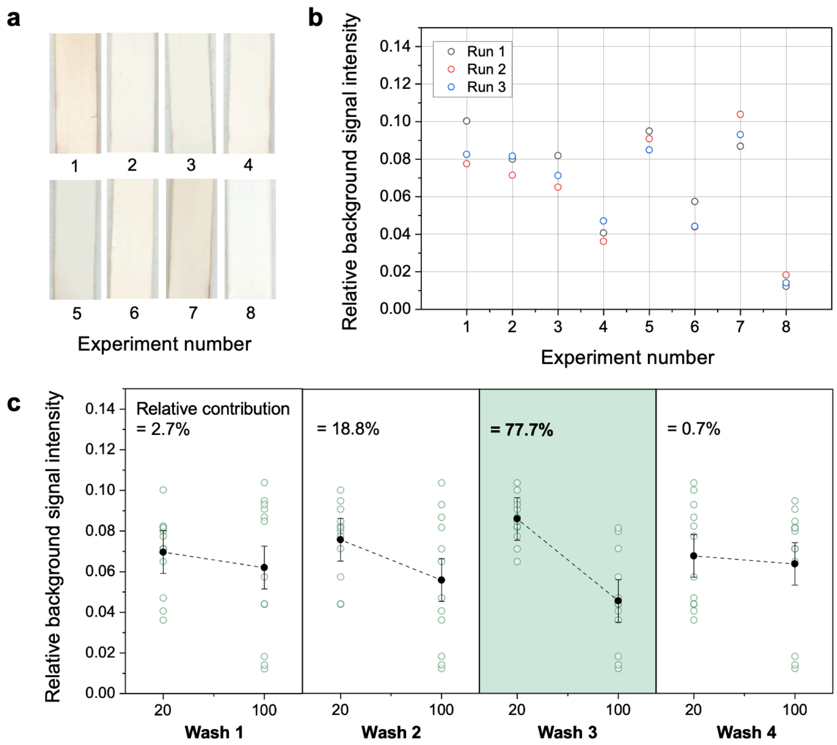

**Figure S3. Relative contribution of each wash step.** (a) Nitrocellulose membranes and (b) the corresponding relative background intensity of the assay test strip with different wash buffer volumes using the Taguchi $L_8(2^4 \times 4^1)$ orthogonal array. (c) Relative contribution of each wash step on the nitrocellulose background, showing that wash 3, i.e., between the enzyme and substrate step, has the greatest contribution to the generation of background signal on the nitrocellulose membrane. Solid data points show the mean ± standard error across grouped runs; hollow data points show the results across all experimental runs. Relative contributions are shown inset.

##### *2.3. Cross-reactivity in MVTs*

We observed visible cross-reactivity between the anti-IFN-α antibody and IFN-ω test line. This interference could be mitigated by reducing the capture probe density of the IFN-ω antigen, which resolved assay interdependency between the two targets but at the expense of reduced analytical sensitivity of the anti-IFN-ω antibody assay.

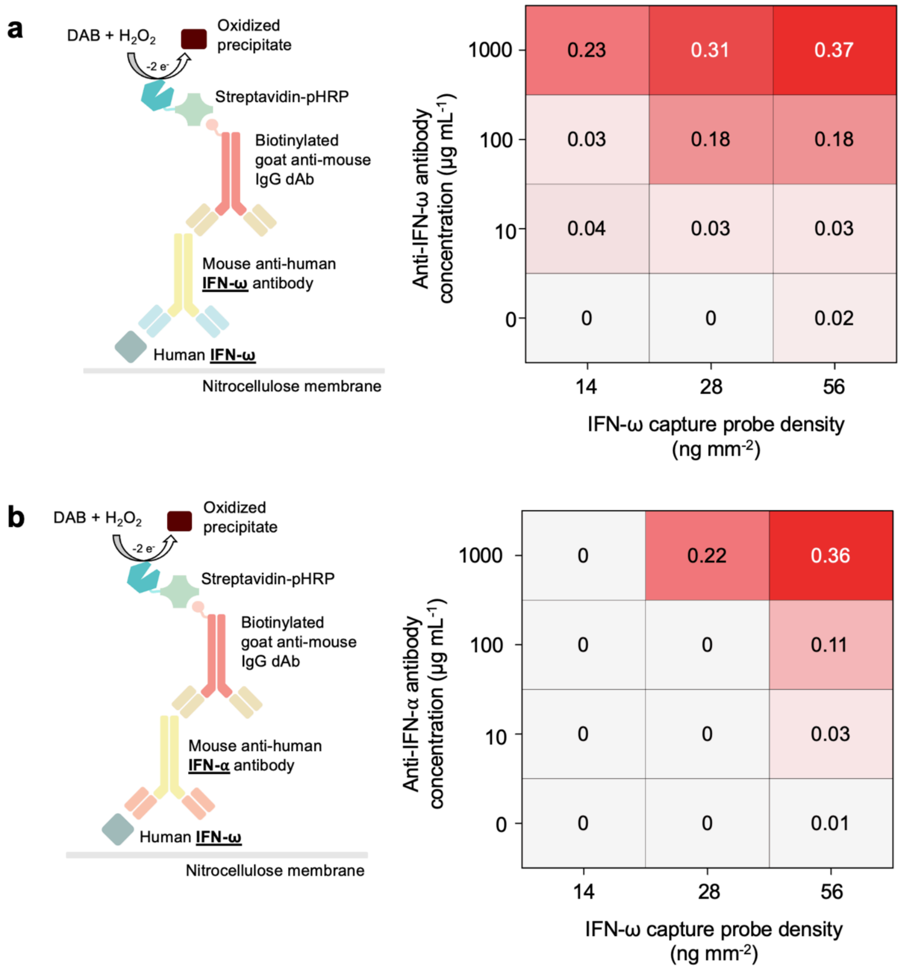

**Figure S4. Cross-reactivity in MVTs.** (a) Specific interaction between the IFN-ω antigen and anti-IFN-ω antibody, and (b) cross-reactivity between the IFN-ω antigen and anti-IFN-α antibody, at varying sample antibody concentrations and capture probe densities. Design of the sandwich immunoassay is schematized on the left and a heatmap showing the relative signal intensity of the nitrocellulose membrane IFN-ω test line is shown on the right.

#### 3. 3D-printed capillaric MVTs

##### *3.1. Logistic fit model, limit of detection, and visual threshold*

Capillaric MVT test line intensities were quantified using ImageJ to obtain the mean colorimetric intensity with subtracted adjacent up- and downstream backgrounds, as previously described.[2] Colorimetric signal intensities were normalized and rescaled from a 0-65,535 16-bit grayscale format to 0-1 relative signal intensities, and plotted as a function of analyte concentration to obtain a binding curve. The curve was fitted to a 4-parameter logistic regression as follows:

$$y = d + \frac{(a-d)}{\left(1+\left(\frac{x}{c}\right)^{b}\right)}$$

where $a$ and $d$ are the theoretical responses at zero and infinite concentration, i.e., minimum and maximum asymptotes of the fit, respectively, $c$ is the inflection point at the middle of the curve, and $b$ is the slope factor (Hill slope) in the linear range.

From the fit, the LOD is calculated as follows:

$$\text{LOD} = \text{mean of blank} + \left(3.3 \times (\text{standard deviation of the blank})\right)$$

where the blank is the sample with zero concentration of analyte.

The visual threshold is defined as the lowest tested concentration at which a signal is discernible and detectable by the naked eye.

#### 4. Identification of key collective parameters governing background suppression

##### *4.1. Potential factors contributing to NSB*

To identify the source of NSB and background on the test lines, we rule out potential causes including cross-reactivity between the antigen test lines and anti-human IgG dAbs, **Figure S5**, and blocking proteins, **Figure S6**. Background intensities were sensitive to plasma dilution, **Figure S7**, suggesting that NSB was driven by plasma constituents rather than the reagents.

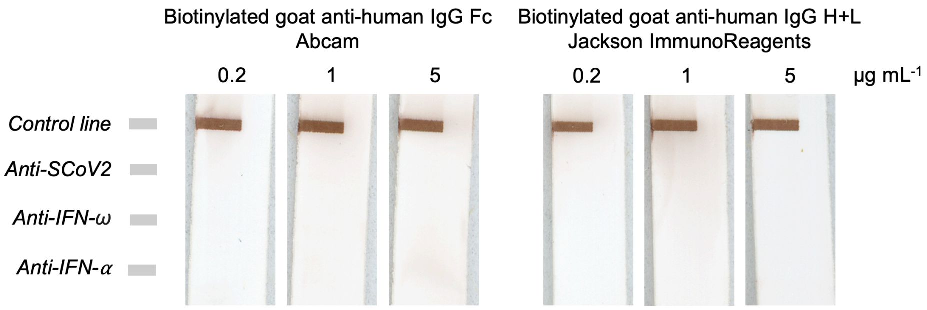

**Figure S5. NSB of anti-human IgG detection antibody with the spotted antigens.** MVTs run with ELISA-buffer as the sample followed by two different biotinylated goat anti-human IgG detection antibodies targeting either the Fc or the H+L regions of the IgG and from different vendors to evaluate whether the dAb contributes to NSB at the test line. For varying concentrations (0.2, 1, and 5 µg $mL^{-1}$) of dAb, there is no visible cross-reactivity with the antigens, so this is not a source of NSB.

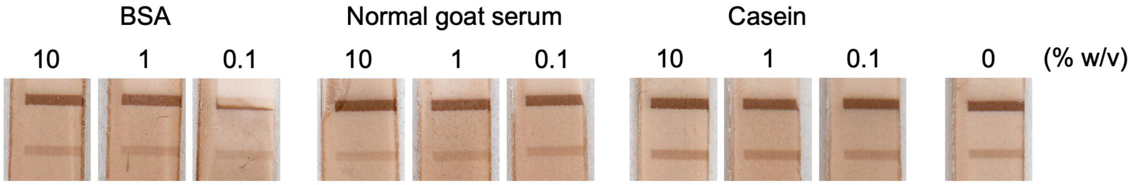

**Figure S6. NSB of blocking proteins.** Nitrocellulose membranes blocked with different proteins including BSA, normal goat serum, and casein do not show changes in the NSB on the IFN-α test line (bottom) or the specific signal of the control line (top) for varying concentrations ranging from 10, 1, and 0.1% (w/v), suggesting that interactions of plasma proteins with blocking proteins used in common ELISA systems does not contribute to NSB, i.e., no anti-BSA or heterophilic antibodies disrupting the response for the anti-IFN-α assay.

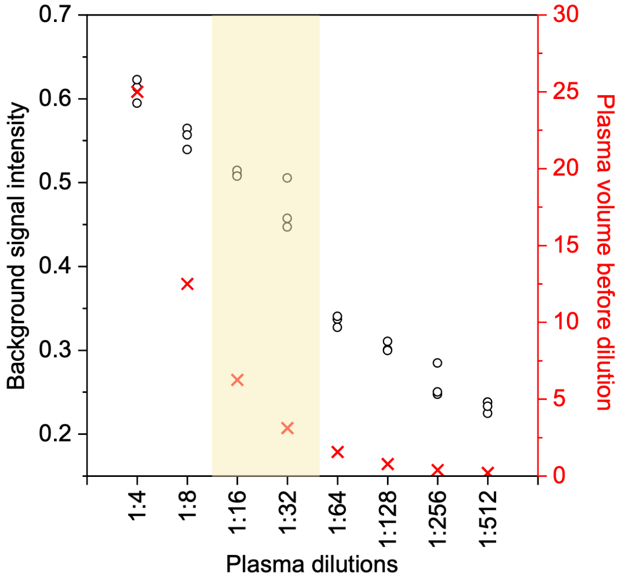

**Figure S7. Selection of plasma dilution for buffer composition DoE.** Background signal intensity decreased with increasing plasma dilution, indicating that NSB was primarily contributed by plasma. Higher dilution (1:64 and higher) resulted in responses near zero that would limit the ability to resolve the effects of buffer composition during DoE, and lower dilution (1:8 and above) led to excessive background signal and high nitrocellulose background that impeded capillary flow. Therefore, 1:16 and 1:32 plasma dilutions were selected as they maintained visually discernible background signals to evaluate and optimize background suppression through buffer formulation while preserving capillary flow through the membrane.

##### *4.2. Diluent buffer composition to supress visual background and quantification of parameter and interaction weights*

Unlike prior DoE rounds, parameters and their ranges for buffer composition that addressed NSB of plasma components to the test line antigens were not well known from prior work for a nitrocellulose membrane assay. In our initial DoE investigating reagent concentrations and volumes (see **Figure 2d**), prior knowledge from ELISA-chips and microplate ELISAs was used to define signal-producing parameter ranges. This ensured sampling of the relevant design space, which is critical in small experimental designs such as the Taguchi $L_{27}(3^{13})$ orthogonal array; otherwise, non-informative ranges could yield zero signal across multiple runs and artificially inflate variance contributions. However, buffer optimization for raAb detection in plasma had not previously been established. Prior ELISA-chip studies relied on standard ELISA buffers (e.g., PBST 0.05%) and were not performed using patient plasma. In contrast, previously reported autoantibody assay buffers were generally developed to minimize NSB on microplate surfaces, were tailored to specific antigens, or consisted of proprietary formulations with undisclosed compositions.[3,4] Consequently, buffer and dilution conditions that achieved sufficient background control while maintaining measurable signal remained to be determined empirically.

We used three DoE rounds with progressively narrower parameter ranges (1) to first identify the relevant parameters and their signal-producing range, and to determine whether buffer composition alone could reduce or eliminate NSB, then (2) to parse parameters and identify NSB-governing factors, and finally (3) to refine the buffer composition and determine whether there are any interactions confounding the experiments.

In the first round of DoE, we used an exploratory Taguchi $L_{18}(2^1 \times 3^7)$ orthogonal array for the anti-IFN-α antibody assay. We examined various buffer constituents varied across 2-3 levels with nested parameters including buffer type and its concentration, surfactant type and its concentration, and NaCl concentration. As the design was exploratory, factors were intentionally not fully decoupled (e.g., pH from buffer type, ionic strength from buffer type, etc.) to first assess the feasibility of using DoE for buffer optimization to mitigate NSB. The conditions were run with 1:16 plasma dilution, **Table S5**. The assay signals are quantified in **Table S6**, and nitrocellulose membranes and response graphs are given in **Figure S8**.

**Table S5. Exploratory Taguchi $L_{18}(2^1 \times 3^7)$ orthogonal array experimental configuration for buffer composition***

| Exp# | Buffer type | Buffer concentration (mM) | Surfactant type | Surfactant concentration (%v/v) | NaCl concentration (mM) |
|---|---|---|---|---|---|
| 1 | Tris | 50 | Tween 20 | 0.25 | 150 |
| 2 | Tris | 250 | CHAPS | 0.5 | 150 |
| 3 | Tris | 1250 | NP-40 | 1 | 150 |
| 4 | Phosphate | 50 | Tween 20 | 0.5 | 150 |
| 5 | Phosphate | 250 | CHAPS | 1 | 150 |
| 6 | Phosphate | 1250 | NP-40 | 0.25 | 150 |
| 7 | Citrate | 50 | CHAPS | 0.25 | 150 |
| 8 | Citrate | 250 | NP-40 | 0.5 | 150 |
| 9 | Citrate | 1250 | Tween 20 | 1 | 150 |
| 10 | Tris | 50 | NP-40 | 1 | 600 |
| 11 | Tris | 250 | Tween 20 | 0.25 | 600 |
| 12 | Tris | 1250 | CHAPS | 0.5 | 600 |
| 13 | Phosphate | 50 | CHAPS | 1 | 600 |
| 14 | Phosphate | 250 | NP-40 | 0.25 | 600 |
| 15 | Phosphate | 1250 | Tween 20 | 0.5 | 600 |
| 16 | Citrate | 50 | NP-40 | 0.5 | 600 |
| 17 | Citrate | 250 | Tween 20 | 1 | 600 |
| 18 | Citrate | 1250 | CHAPS | 0.25 | 600 |

*Columns 6-8 are empty and used for assessing parameter interactions, so while they do not encode an experimental condition, they are included in the design for extra degrees of freedom.

**Table S6. ANOVA, effect size, and percent relative contribution of assay parameters for buffer composition**

| Factor | Degree of freedom | Sum of squares | F ratio | $\eta^2$ effect size [0-1] | Relative contribution (%) |
|---|---|---|---|---|---|
| **Buffer type** | 2 | 0.00257 | 2.6 | 0.12 | 15.5 |
| **Buffer concentration (mM)** | 1 | 0.000192 | 0.4 | 0.009 | 1.2 |
| **Surfactant type** | 2 | 0.0139 | 13.9 | 0.64 | 83.3 |
| **Surfactant concentration (%v/v)** | 1 | 1.53e-5 | 0.03 | 0.0007 | 0.09 |
| **NaCl concentration (mM)** | 1 | 2.02e-6 | 0.004 | 0.0001 | 0.01 |

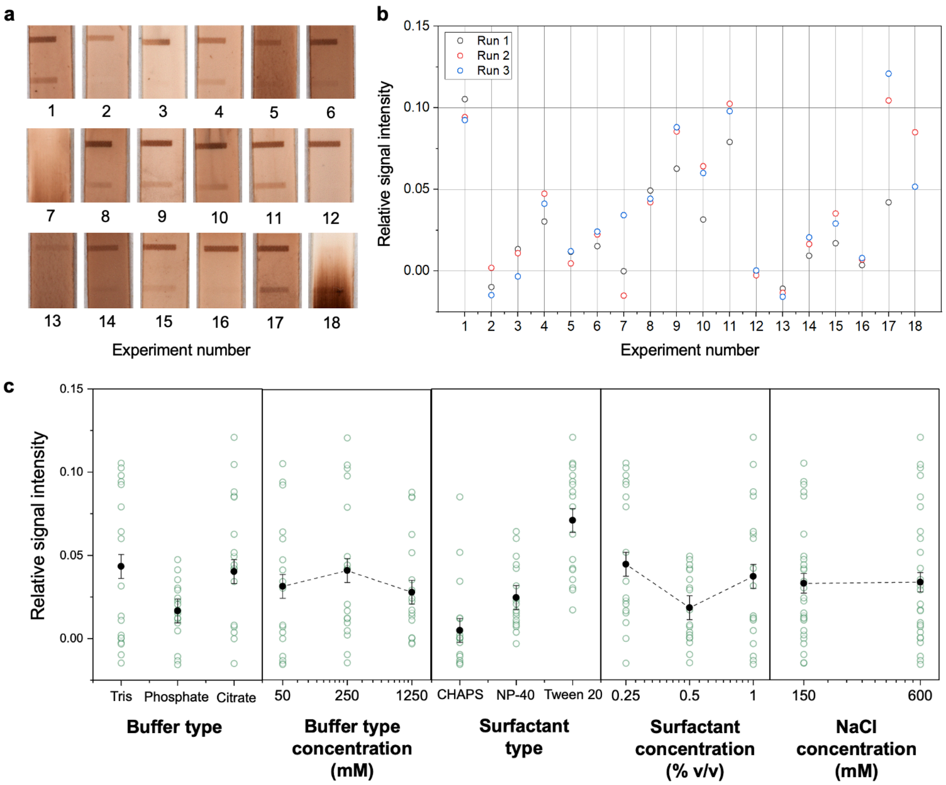

**Figure S8. Buffer composition exploration using Taguchi $L_{18}(2^1 \times 3^7)$ orthogonal array results.** (a) Nitrocellulose membranes with test lines (bottom) and control lines (top) and (b) their corresponding IFN-α relative test line signal intensity with different buffer compositions using the Taguchi $L_{18}(2^1 \times 3^7)$ orthogonal array. (c) Response graphs showing the contribution of individual buffer components on the relative signal intensity of the test line. Solid data points show the mean ± standard error across grouped runs; hollow data points show the results across all experimental runs.

To parse the contribution of buffer composition on NSB, a Taguchi $L_{32}(2^1 \times 4^9)$ orthogonal array was run at two plasma dilutions including 1:32 and 1:16 plasma:buffer. Factors were parsed by keeping the buffer type at a fixed concentration (10 mM) to avoid experimentally confounding with ionic strength or pH. The parsed buffer DoE included buffer type, pH, ionic strength, surfactant type, and surfactant concentration, **Table S7**. The nitrocellulose membranes and response graphs for each buffer is given in **Figure 4c** and **Figure S9-10**, and the relative contribution quantified in **Table S8**.

**Table S7. Buffer composition and optimization using Taguchi $L_{32}(2^1 \times 4^9)$ orthogonal array experimental configuration***

| Exp# | Buffer type | pH | Ionic strength (mM) | Surfactant type | Surfactant concentration (%v/v) |
|---|---|---|---|---|---|
| 1 | Tris | 6 | 50 | Tween 20 | 0.01 |
| 2 | Tris | 6 | 150 | Brij-35 | 0.05 |
| 3 | Tris | 6 | 450 | NP-40 | 0.1 |
| 4 | Tris | 6 | 1350 | Triton-X-100 | 1 |
| 5 | Tris | 7 | 50 | Tween 20 | 0.05 |
| 6 | Tris | 7 | 150 | Brij-35 | 0.01 |
| 7 | Tris | 7 | 450 | NP-40 | 1 |
| 8 | Tris | 7 | 1350 | Triton-X-100 | 0.1 |
| 9 | Tris | 8 | 50 | Brij-35 | 0.1 |
| 10 | Tris | 8 | 150 | Tween 20 | 1 |
| 11 | Tris | 8 | 450 | Triton-X-100 | 0.01 |
| 12 | Tris | 8 | 1350 | NP-40 | 0.05 |
| 13 | Tris | 9 | 50 | Brij-35 | 1 |
| 14 | Tris | 9 | 150 | Tween 20 | 0.1 |
| 15 | Tris | 9 | 450 | Triton-X-100 | 0.05 |
| 16 | Tris | 9 | 1350 | NP-40 | 0.01 |
| 17 | PBS | 6 | 50 | Triton-X-100 | 0.01 |
| 18 | PBS | 6 | 150 | NP-40 | 0.05 |
| 19 | PBS | 6 | 450 | Brij-35 | 0.1 |
| 20 | PBS | 6 | 1350 | Tween 20 | 1 |
| 21 | PBS | 7 | 50 | Triton-X-100 | 0.05 |
| 22 | PBS | 7 | 150 | NP-40 | 0.01 |
| 23 | PBS | 7 | 450 | Brij-35 | 1 |
| 24 | PBS | 7 | 1350 | Tween 20 | 0.1 |
| 25 | PBS | 8 | 50 | NP-40 | 0.1 |
| 26 | PBS | 8 | 150 | Triton-X-100 | 1 |
| 27 | PBS | 8 | 450 | Tween 20 | 0.01 |
| 28 | PBS | 8 | 1350 | Brij-35 | 0.05 |
| 29 | PBS | 9 | 50 | NP-40 | 1 |
| 30 | PBS | 9 | 150 | Triton-X-100 | 0.1 |
| 31 | PBS | 9 | 450 | Tween 20 | 0.05 |

| | | | | | |
|---|---|---|---|---|---|
| **32** | PBS | 9 | 1350 | Brij-35 | 0.01 |

*Columns 6-10 are empty and used for assessing parameter interactions, so while they do not encode an experimental condition, they are included in the design for extra degrees of freedom.

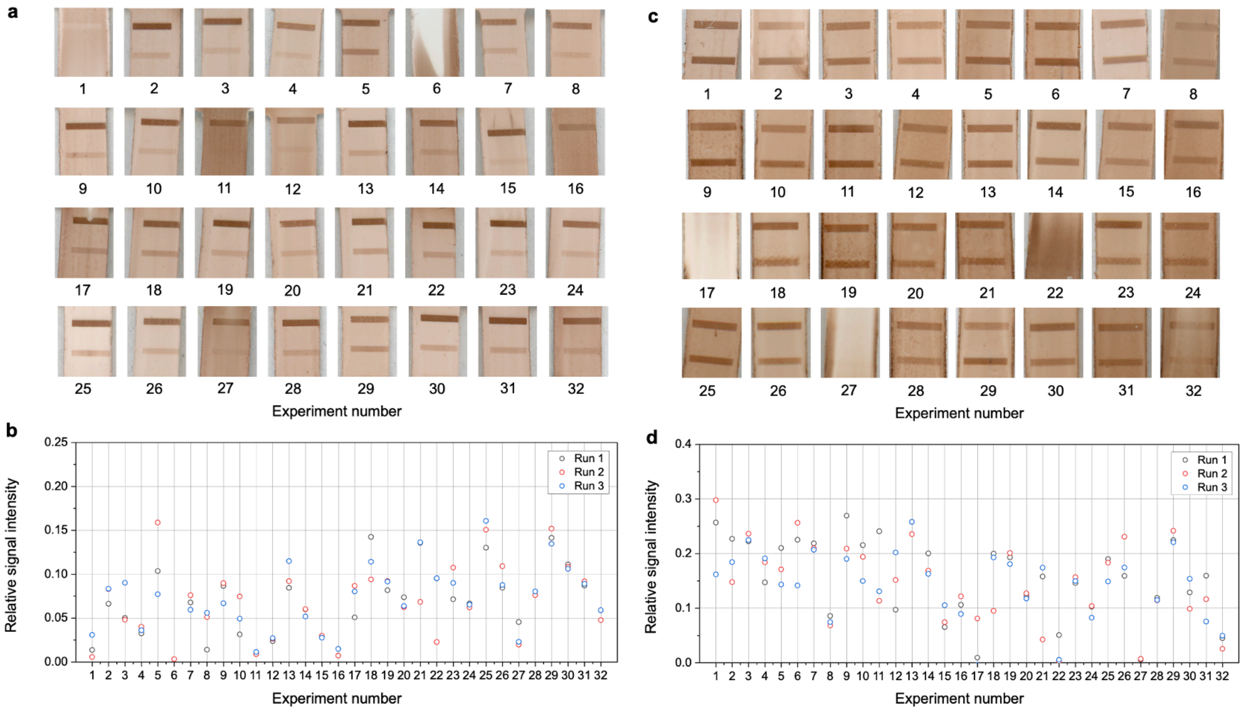

**Figure S9. Buffer composition and optimization using Taguchi $L_{32}(2^1 \times 4^9)$ orthogonal array results.** (a, c) Nitrocellulose membranes and (b, d) the corresponding relative signal intensity of the IFN-α test line (bottom) and control line (top) with different buffer compositions using the Taguchi $L_{32}(2^1 \times 4^9)$ orthogonal array at both 1:32 (left) and 1:16 plasma dilutions (right).

**Table S8. ANOVA, effect size, and percent relative contribution of assay parameters and parameter interactions for buffer composition and optimization**

| **1:32 plasma:buffer dilution** | | | | | |
|---|---|---|---|---|---|
| **Factor** | **Degree of freedom** | **Sum of squares** | **F ratio** | **$\eta^2$ effect size [0-1]** | **Relative contribution (%)** |
| **Buffer type** | 1 | 0.0140 | 32.6 | 0.39 | 55.4 |
| **pH** | 1 | 0.000164 | 0.4 | 0.005 | 0.7 |
| **Ionic strength (mM)** | 1 | 0.00713 | 16.6 | 0.20 | 28.2 |
| **Surfactant type** | 3 | 0.00251 | 1.9 | 0.07 | 9.9 |
| **Surfactant concentration (%v/v)** | 1 | 0.00148 | 3.4 | 0.04 | 5.8 |
| **1:16 plasma:buffer dilution** | | | | | |
| **Factor** | **Degree of freedom** | **Sum of squares** | **F ratio** | **$\eta^2$ effect size [0-1]** | **Relative contribution (%)** |
| **Buffer type** | 1 | 0.00588 | 5.7 | 0.07 | 9.3 |
| **pH** | 1 | 0.00774 | 7.6 | 0.09 | 12.2 |
| **Ionic strength (mM)** | 1 | 0.0331 | 32.4 | 0.37 | 52.1 |
| **Surfactant type** | 3 | 0.0111 | 3.6 | 0.13 | 17.5 |
| **Surfactant concentration (%v/v)** | 1 | 0.00567 | 5.6 | 0.06 | 8.9 |

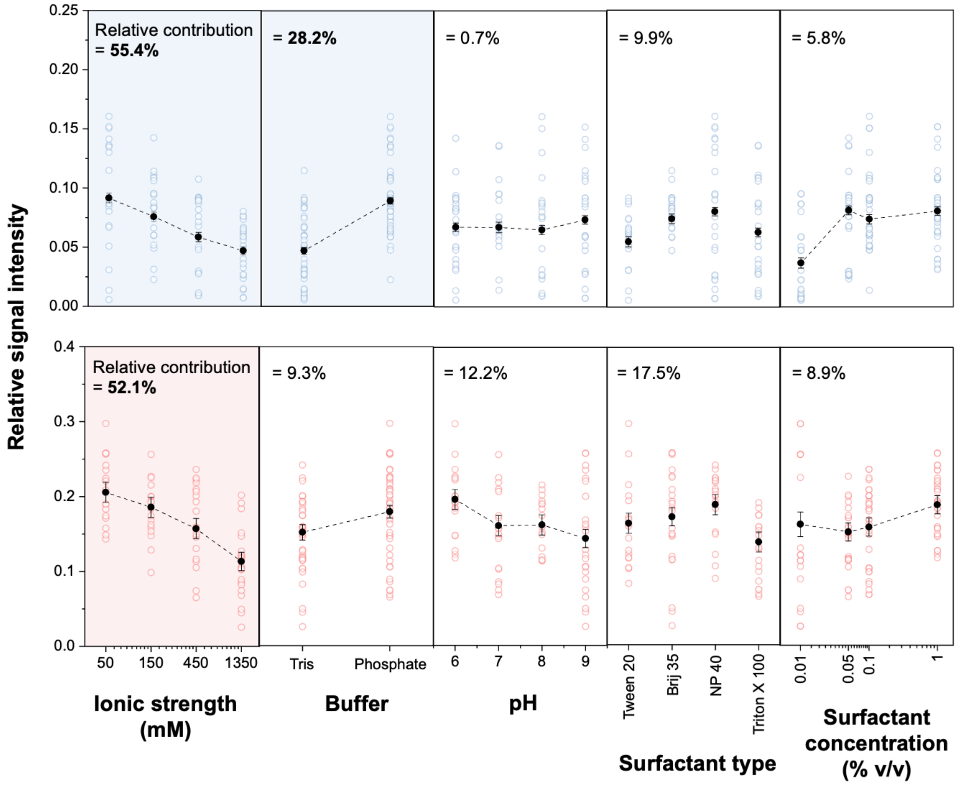

**Figure S10. Response graphs of buffer composition and optimization Taguchi $L_{32}(2^1 \times 4^9)$ orthogonal array.** Scatter plots illustrating the effect of each parameter on assay signal; insets show the corresponding quantified relative signal intensities. Responses at both 1:32 plasma dilution (top, blue) and 1:16 plasma dilution (bottom, red) are shown. Solid data points show the mean ± standard error across grouped runs; hollow data points show the results across all experimental runs. Relative contributions are shown inset.

We investigated parameter interactions on a smaller DoE with narrow parameter ranges parse parameter confounding. This was done with a Taguchi $L_9(3^4)$ orthogonal array by varying the Tris, Tween 20, and NaCl concentrations at three levels. There are three two-way interactions for this design: Tris × NaCl concentrations, NaCl × Tween 20 concentrations, and Tween 20 × Tris concentrations. With only 9 experiments, 8 degrees of freedom were available and 3 were assigned to the continuous parameters, leaving behind 5 for interactions (each requiring 2) and error. Thus, the Taguchi $L_9(3^4)$ orthogonal array with 3 continuous parameters is biased towards main effects, thus the effect size of a two-way interaction will always be lower than individual main effects, so ANOVA could not be used to identify or quantify interactions. Instead, parameters were plotted against each other to determine whether a given parameter response is conditional on another. In this case, changes in the slope direction and non-flat responses would indicate the presence of conditional dependence between parameters. The DoE experimental configuration is given in **Table S9**, quantified parameter weights in **Table S10**, nitrocellulose membranes, quantitative readouts, and response graphs in **Figure S11**, and conditional mean plots in **Figure S12**.

**Table S9. Identifying 2-way interactions using Taguchi $L_9(3^4)$ orthogonal array experimental configuration***

| Exp# | Tris concentration (mM) | Tween 20 concentration (%v/v) | NaCl concentration (mM) |
|---|---|---|---|
| **1** | 50 | 0.01 | 37.5 |
| **2** | 50 | 0.1 | 150 |
| **3** | 50 | 1 | 600 |
| **4** | 250 | 0.01 | 150 |
| **5** | 250 | 0.1 | 600 |
| **6** | 250 | 1 | 37.5 |
| **7** | 1250 | 0.01 | 600 |
| **8** | 1250 | 0.1 | 37.5 |
| **9** | 1250 | 1 | 150 |

*Column 4 is empty and used for assessing parameter interactions, so while it does not encode an experimental condition, it is included in the design for extra degrees of freedom.

**Table S10. ANOVA, effect size, and percent relative contribution of assay parameters and parameter interactions for identifying 2-way interactions**

| Factor | Degree of freedom | Sum of squares | F ratio | $\eta^2$ effect size [0-1] | Relative contribution (%) |
|---|---|---|---|---|---|
| **Tris concentration (mM)** | 1 | 0.0924 | 9.7 | 0.49 | 65.3 |
| **Tween 20 concentration (%v/v)** | 1 | 0.0255 | 2.7 | 0.13 | 18.0 |
| **NaCl concentration (mM)** | 1 | 0.0236 | 2.5 | 0.12 | 16.6 |

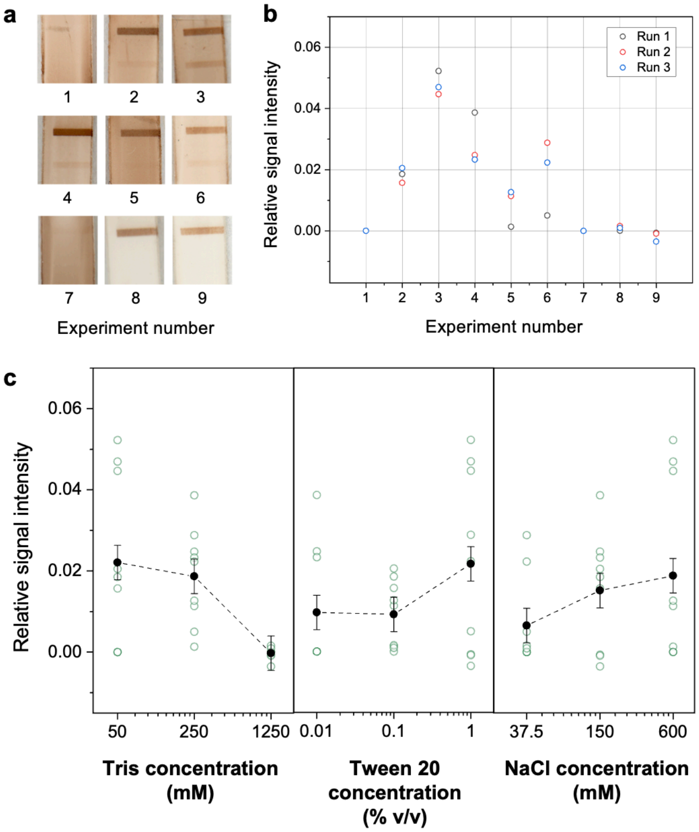

**Figure S11. Identifying 2-way interactions using Taguchi $L_9(3^4)$ orthogonal array results.** (a) Nitrocellulose membranes with IFN-α test line (bottom) and control line (top) and (b) the corresponding test line relative signal intensity with different buffer compositions using the Taguchi $L_9(3^4)$ orthogonal array. (c) Response graphs showing the contribution of individual buffer components on the relative signal intensity. Solid data points show the mean ± standard error across grouped runs; hollow data points show the results across all experimental runs.

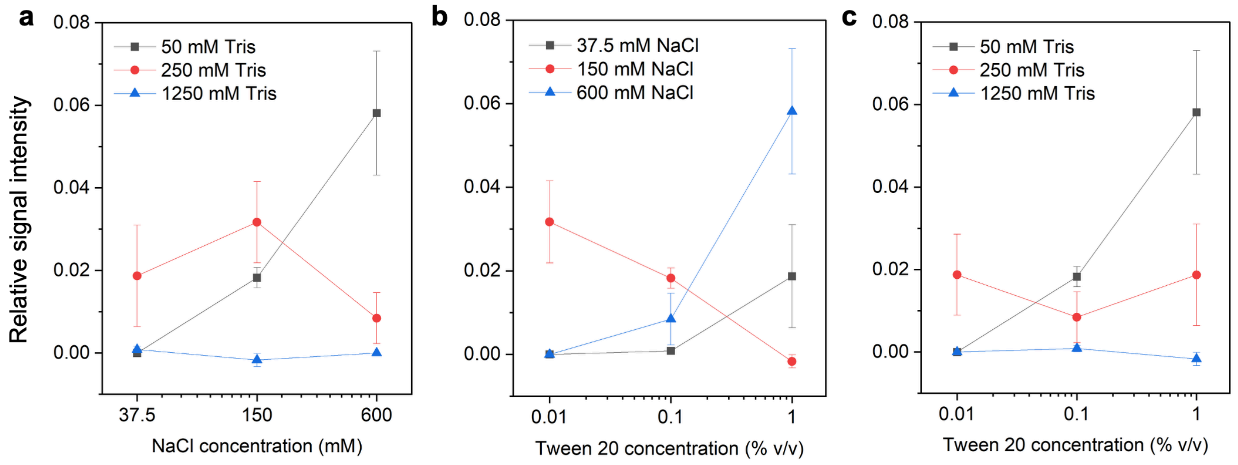

**Figure S12. Conditional mean plots for 2-way interactions** for (a) NaCl × Tris concentrations, (b) Tween 20 × NaCl concentrations, and (c) Tween 20 × Tris concentrations. Data points show mean ± standard error across grouped runs.

From the conditional mean plots, Tris × NaCl concentration interaction was identified with an increasing positive slope for low Tris, a reversal for mid-Tris, and flattening for high Tris concentrations as a function of NaCl concentration, indicating the presence of a conditional relationship between the parameters. Interestingly, a reversal in slope was also seen for NaCl and Tween 20 interactions, but this was largely given by the point where there is 150 mM NaCl and 1% (v/v) Tween 20 (experiment 9), which had high Tris concentration, so we speculate possible bleed-through from the Tris × NaCl interaction could be confounding this point. Tris × Tween 20 concentration are conditional at low Tris concentration, then the response flattened at higher concentrations, suggesting that at higher salt concentrations, Tween 20 concentration is independent of other parameters. Therefore, a response given by the DoE could be either attributed to increasing Tris or decreasing NaCl concentration; however, the factors are not completely independent, so these parameters need to be adjusted together.

##### *4.3. Surface tension of buffer components*

**Table S11. Surface tension of buffer components**

| **Composition** | **Concentration** | **Surface tension (mN $m^{-1}$)** |
|---|---|---|
| MilliQ water | | 72 |
| NaCl in MilliQ water | 50 mM | 72 |
| | 150 mM | 72 |
| | 450 mM | 72 |
| | 750 mM | 70 |
| | 1350 mM | 71 |
| Tris base in MilliQ water | 10 mM | 72 |
| | 50 mM | 72 |
| | 250 mM | 72 |
| | 500 mM | 71 |
| | 750 mM | 72 |
| | 1250 mM | 72 |
| Goat serum in MilliQ water | 0.1% (v/v) | 63 |
| | 1% (v/v) | 55 |
| | 10% (v/v) | 51 |
| Tween 20 in MilliQ water | 0.01% (v/v) | 41 |
| | 0.05% (v/v) | 39 |
| | 0.1% (v/v) | 38 |
| | 1% (v/v) | 37 |

**Table S12. Surface tension of select non-ionic surfactants**

| **Composition** | **Concentration** | **Surface tension (mN $m^{-1}$)** |
|---|---|---|
| Brij 35 in MilliQ water | 0.01% (v/v) | 46 |
| | 0.05% (v/v) | 40 |
| | 0.1% (v/v) | 40 |
| | 1% (v/v) | 40 |
| NP-40 in MilliQ water | 0.01% (v/v) | 31 |
| | 0.05% (v/v) | 31 |

| | | |
|---|---|---|
| | 0.1% (v/v) | 31 |
| | 1% (v/v) | 31 |
| Triton X-100 in MilliQ water | 0.01% (v/v) | 37 |
| | 0.05% (v/v) | 31 |
| | 0.1% (v/v) | 31 |
| | 1% (v/v) | 31 |

**Table S13. Surface tension of buffer compositions**

| **Composition** | **Surface tension (mN $m^{-1}$)** |
|---|---|
| 500 mM tris base + 500 mM NaCl in MilliQ water | 72 |
| 500 mM tris base + 500 mM NaCl + 5% (v/v) goat serum in MilliQ water | 53 |
| 500 mM tris base + 500 mM NaCl + 5% (v/v) goat serum + 0.01% (v/v) tween 20 in MilliQ water | 46 |

#### 5. Benchmarking and clinical assessment of capillaric MVTs

##### *5.1. Patient cohort information*

**Table S14. Patient cohort information**

| ID | Sex | Age range* | Year of first infection**, *** | Time from infection to blood draw**,**** | Infection intensity** | Outcome | Additional notes on infection severity |
|---|---|---|---|---|---|---|---|
| 1 | F | 86-90 | 2020 | 16 months | 3 Moderate Illness | Survived | |
| 2 | M | 81-85 | 2022 | 4 months | 2 Mild Illness | Survived | |
| 3 | F | 81-85 | 2020 | 16 months | 1 Asymptomatic or Presymptomatic Infection | Survived | |
| 4 | M | 76-80 | 2022 | 1 month | 2 Mild Illness | Survived | |
| 5 | F | 81-85 | 2020 | 16 months | 1 Asymptomatic or Presymptomatic Infection | Survived | |
| 6 | F | 71-75 | 2020 | 4 months | 2 Mild Illness | Survived | |
| 7 | M | 61-65 | 2020 | 5 months | 1 Asymptomatic or Presymptomatic Infection | Survived | |
| 8 | M | 21-25 | NA | NA | 0 Unknown | Survived | |
| 9 | M | 31-35 | NA | NA | 0 Unknown | Survived | |
| 10 | M | NA | NA | NA | 0 Unknown | Survived | |
| 11 | M | 26-30 | NA | NA | 0 Unknown | Survived | |
| 12 | F | 86-90 | 2020 | <1 month | 5 Critical Illness | Deceased | Required supplementary oxygen |
| 13 | F | 81-85 | 2020 | 1 month | 1 Asymptomatic or Presymptomatic Infection | Survived | |
| 14 | F | 96-100 | NEVER | NA | NA | Survived | |
| 15 | M | 81-85 | 2020 | 11 months | 2 Mild Illness | Survived | |

| | | | | | | | |
|---|---|---|---|---|---|---|---|
| 16 | M | 66-70 | NEVER | NA | NA | Survived | |
| 17 | M | 61-65 | 2020 | 10 months | 2 Mild Illness | Survived | |
| 18 | F | 71-75 | NEVER | NA | NA | Survived | |
| 19 | F | 71-75 | NEVER | NA | NA | Survived | |
| 20 | F | 81-85 | 2020 | 10 months | 4 Severe Illness | Survived | |
| 21 | M | 91-95 | 2022 | 11 months | 2 Mild Illness | Survived | |
| 22 | M | 21-25 | NA | NA | 0 Unknown | Survived | |
| 23 | F | 26-30 | NA | NA | 0 Unknown | Survived | |
| 24 | F | 21-25 | NA | NA | 0 Unknown | Survived | |
| 25 | M | NA | NA | NA | 0 Unknown | Survived | |
| 26 | F | 26-30 | NA | NA | 0 Unknown | Survived | |
| 27 | M | 26-30 | NA | NA | 0 Unknown | Survived | |
| 28 | M | 76-80 | 2020 | <1 month | 4 Severe Illness | Survived | Required supplementary oxygen |
| 29 | M | 81-85 | 2020 | <1 month | 5 Critical Illness | Deceased | Required supplementary oxygen |
| 30 | M | 91-95 | 2020 | 1 month | 5 Critical Illness | Survived | ICU admitted, required supplementary oxygen |
| 31 | M | 81-85 | 2020 | <1 month | 4 Severe Illness | Survived | |
| 32 | F | 81-85 | 2021 | <1 month | 5 Critical Illness | Deceased | ICU admitted, required supplementary oxygen |
| 33 | F | 81-85 | 2020 | <1 month | 4 Severe Illness | Survived | ICU admitted, required supplementary oxygen |
| 34 | M | 66-70 | 2020 | 1 month | 4 Severe Illness | Survived | Required supplementary oxygen |
| 35 | M | 91-95 | 2020 | <1 month | 5 Critical Illness | Deceased | Required supplementary oxygen |
| 36 | F | 86-90 | 2020 | <1 month | 5 Critical Illness | Deceased | Required supplementary oxygen |
| 37 | F | NA | NEVER | NA | NA | NA | |
| 38 | F | NA | NEVER | NA | NA | NA | |

| | | | | | | |
|---|---|---|---|---|---|---|
| 39 | M | NA | NEVER | NA | NA | NA |
| 40 | M | NA | NEVER | NA | NA | NA |
| 41 | M | NA | NEVER | NA | NA | NA |

* 5-year non-overlapping age ranges, e.g., 21-25, 26-30, 31-35, 36-40, and so on.

** NEVER = indicates no prior COVID-19 infection, NA = information not available.

*** Tested for infection using standard laboratory PCR.

**** IDs 37-41 are pre-pandemic controls with blood drawn between 2016-2018.

##### *5.2. Pre-antigen-bound raAbs in circulation*

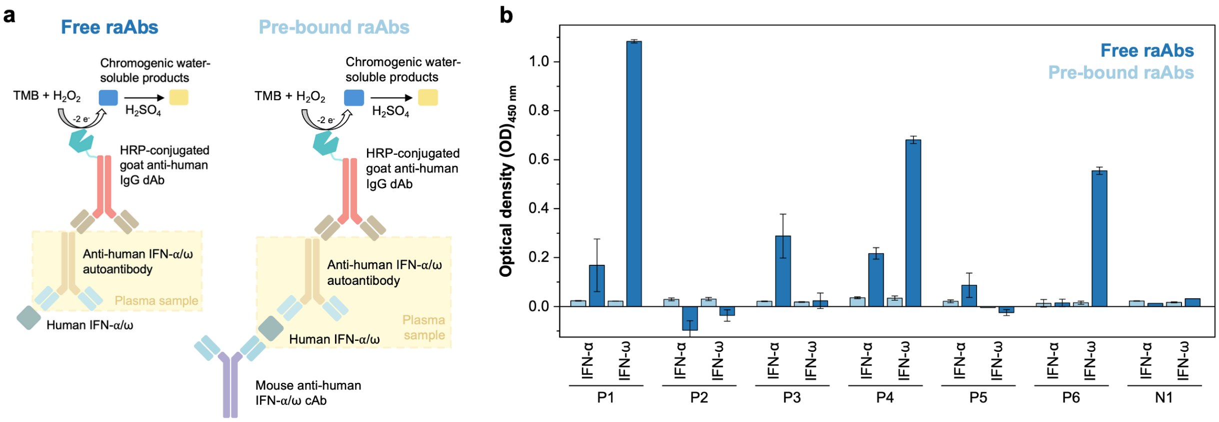

**Figure S13. Impact of anti-IFN-α and -ω raAbs in plasma pre-bound to their antigen.** (a) Microplate ELISA design to detect free (left) and antigen-pre-bound (right) raAbs in plasma, evaluating whether pre-bound raAbs influence assay performance and whether they are present at appreciable levels in patient samples. (b) Microplate ELISA results for six representative patient plasma samples (P1-P6), including both anti-IFN-α- and anti-IFN-ω-positive and -negative samples, together with a pre-pandemic control (N1), showing that antigen-pre-bound raAbs are less abundant than their free counterparts. Bars show the mean ± standard deviation across triplicate microplate ELISA wells.

##### *5.3. Empirical rounds to synchronize visual and clinical thresholds on capillaric MVTs*

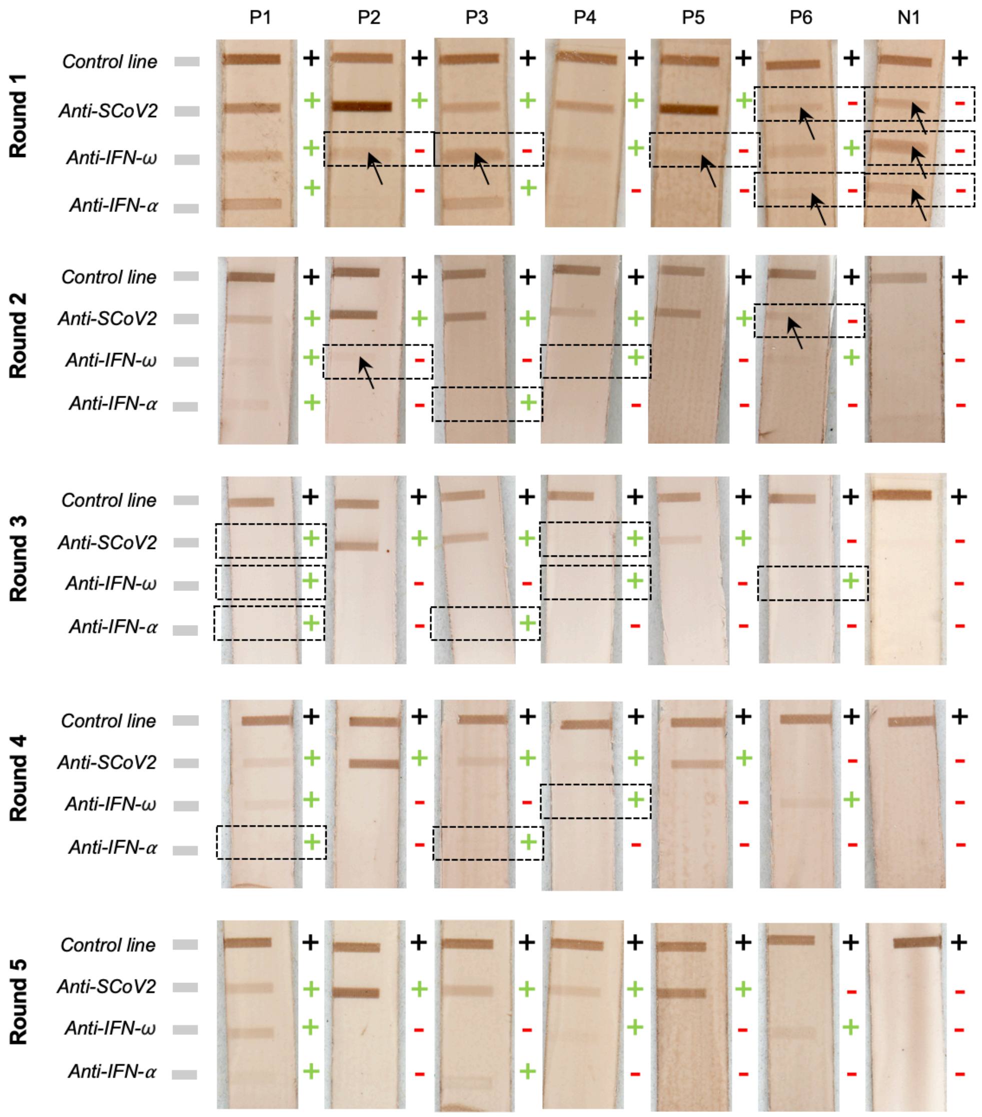

**Figure S14. Testing and optimizing the visual threshold of capillaric MVTs for synchronization with clinical positivity threshold.** Visual readout optimization of capillaric MVTs was performed with DoE-guided parameter tuning to first collectively reduce the background and then individually adjust capture probe density to reach the clinical threshold. Nitrocellulose membranes show the visual readout of six positive patient controls (P1-6) and one pre-pandemic negative control (N1). Black + symbols show the presence of a control line, and coloured +/- symbols show the expected result of the control plasma samples based on microplate ELISA as the reference method. Test lines in dashed black boxes show the presence of false readout with arrows identifying the visible test line. We started with a high salt buffer that led to false positives across all assays (round 1); then Tris concentration was progressively increased while keeping NaCl lower, resulting in false negatives (rounds 2-3). Then, we again reduced Tris, while also increasing the capture probe density of IFN-α and -ω to restore the test line signals and align with the microplate ELISA readouts (rounds 4-5).

##### *5.4. Capillaric MVT readouts of clinical samples*

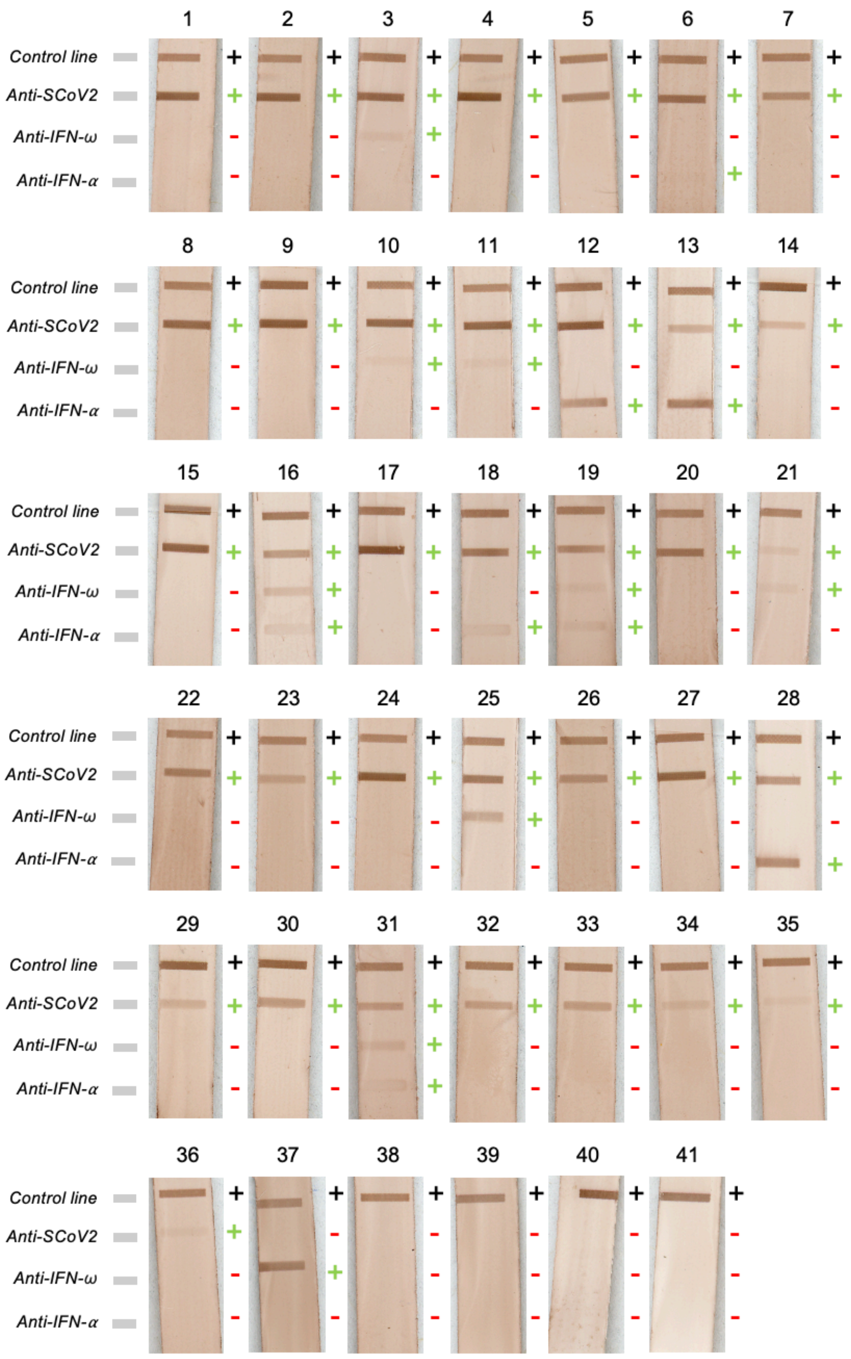

**Figure S15. Capillaric MVT readouts of clinical samples.** Capillaric MVT readouts of 41 clinical plasma samples with assays starting from the bottom, including the anti-IFN-α, anti-IFN-ω, and anti-SCoV2 test lines, and the BSA-biotin control line. Black + symbols show the presence of a control line, and coloured +/- symbols show the expected result of the control plasma samples based on microplate ELISA as the reference method.

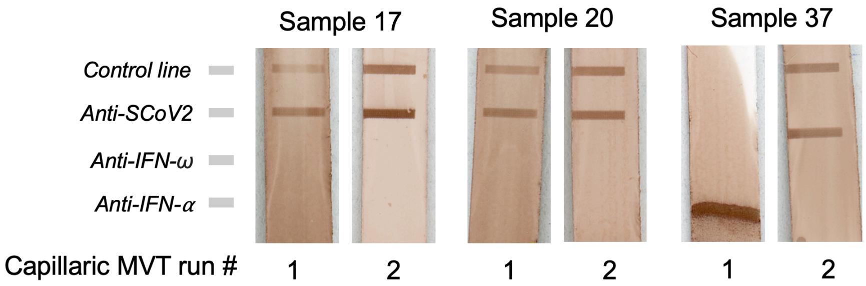

**Figure S16. Capillaric MVT yield.** Clinical plasma samples were run once on every 3D-printed capillaric MVT, except for three instances where readouts that were not interpretable, i.e., absence or faint control line, which prompted running the sample on a second chip. Failed runs were attributed to poor running of the 3D-printed chips, either due to printing defects or poor chip preparation, i.e., not fully dried after printing, resulting in crosstalk between reservoirs and weak assay readouts (samples 17 and 20), or poor chip-to-assay connection (sample 37) resulting in poor liquid rise over the nitrocellulose membrane and accumulation of the precipitate only at the membrane front. Thus, the yield of successful capillaric MVT operation is 38/41, assembled and operated by a trained user.
